# Factors associated with exclusive breastfeeding among mothers of children under two years of age in Dalit community, Rajbiraj Municipality, Saptari, Nepal

**DOI:** 10.1101/2023.08.09.23292718

**Authors:** Neha Kumari Das, Nirmal Duwadi, Ramchandra Sinha, Alisha Dahal

**Affiliations:** Department of Public Health, Nobel College, Pokhara University, Kathmandu, Bagmati Province, Nepal

## Abstract

The prevalence of exclusive breastfeeding (EBF) is suboptimal in Nepal and very low in Madhesh province. Dalits are commonly recognized for experiencing economic exploitation, a lack of political representation, social marginalization, educational disadvantage, being classified as untouchables, and enduring the denial of basic human dignity and social justice.

Objective of this study is to assess the proportion of mothers of children under 2 years of age, practicing exclusive breastfeeding and associated factors in the Dalit community of Rajbiraj Municipality, Saptari, Nepal.

The study utilized an analytical, cross-sectional design by using a semi-structured questionnaire to 156 Dalit mothers of children under 2 years of age in Rajbiraj Municipality, Saptari, Nepal. In the bivariate analysis, which focuses on exploring the connection between independent variables and dependent variables, chi-squire statistics were utilized. Subsequently, the independent variables that displayed significance in the bivariate analysis were included in the multivariate logistic regression.

The estimated prevalence of exclusive breastfeeding in the population was 43.6%. Mother’s occupation (adjusted odds ratio (AOR = 4.459; CI = 1.444 -13.767), smoking habit (AOR = 2.755; CI = 1.120 – 6.774), colostrum milk feeding (AOR = 12.472; CI = 3.253 – 47.823), number of times visit the health center for ANC (AOR = 2.333; CI = 1.040 – 5.233) were positively associated with exclusive breastfeeding, whereas, sex of the child, type of family, knowledge about breastfeeding, counselling on EBF in ANC and/or PNC visit were also associated with exclusive breastfeeding. Among the respondents who did not practice exclusive breastfeeding, the common reasons cited were trouble initiating milk flow (23.7%), insufficient breast milk production (21.7%), breast milk not satisfying the baby (20.9%), domestic work burden (12%), difficulties with infant sucking or latching (6.4%), and the baby being unable to be breastfed owing to sickness (4.8%).

Maternal education, occupation, colostrum milk feeding, and antenatal care visits were identified as important influencers of exclusive breastfeeding. Healthcare professionals, policymakers, and community leaders can utilize these insights to formulate effective strategies and design intervention that encourage and support exclusive breastfeeding among Dalit mothers, ultimately improving the health and well-being of the infants of the marginalized community.

## Introduction

### Background

Nepal is a nation characterized by rich sociocultural diversity, is spread across seven provinces and three ecological regions: mountain, hills and Terai. Within this diverse tapestry, Nepal is home to 23 distinct caste and ethnic groups.(1)

However, amidst this diversity, one group, the Dalits faces significant challenges. In the hierarchical Hindu caste system, Dalits often endure economic exploitation, political marginalization, social degradation, educational disadvantages, and stigma of untouchables. These injustices compound to deny them basic human dignity and social justice, leading to a life marked by limited income opportunities, reliance on daily wages, livestock sales, and social security allowances. Despite constituting 13.6% of the population, Dalits face formidable barriers to achieving an equitable and prosperous life. (2, 3)

Despite ongoing social disparities, it’s crucial to grasp the vital importance of breastfeeding—an inclusive practice with profound implications for infant health. Breast milk, often hailed as the healthiest nourishment for infants, stands as a precious gift from mothers, meeting all the energy and nutritional needs of a baby.(4) Recognizing the immense value of breastfeeding, the World Health Organization (WHO) advocates for exclusive breastfeeding, wherein infants receive only breast milk during the first six months of life, without any additional food or liquids. (5) Exception Only oral rehydration solution, authorized drugs, vitamins, and minerals for exclusively breastfed infants. (6) Exclusive breastfeeding (EBF) plays a vital role in reducing child morbidity and mortality, as well as contributing to the effective management of healthcare expenses in society. It serves as a significant public health strategy to improve the health of both mothers and children. (7)

The benefits of EBF are far – reaching, encompassing significant protection against various diseases, including infections, pneumonia, sudden infant death syndrome, malocclusion, diarrhea, diabetes, asthma, heart disease, and obesity. It significantly reduce the risk of infant mortality due to infections in the first two years of life. (8) Additionally, EBF supports healthy brain development and is associated with better intelligence tests results in children and adolescents. (5) It fosters a close bond between women and their children while reducing the risk of breast and ovarian cancer. (9) Despite these compelling benefits, the prevalence of exclusive breastfeeding in Nepal remains alarming low. (10–12)

To promote a child’s survival, growth, and development, it is crucial to follow the right feeding and care practices. According to the annual report, the percentage of exclusively breastfeeding infants in 2021 was 36.9%, which was significantly lower than the NDHS 2022 figure of 56.4% and the most recent MICS 2019 figure of 62.1%. According to a breakdown by province, Madhesh Province has the lowest percentage of infants aged 0 to 6 months who are exclusively breastfed, at 26%. Vulnerable communities, such as Dalits, are the least likely to exclusively breastfeed, and rates of exclusive breastfeeding fall far short of the national target due to humiliation and discrimination. (10) The Dalits of Nepal are the most disadvantaged caste group and have benefited least from the advances in maternal and child health services. (13) For example, ANMs, who are front-line healthcare providers from non-Dalit castes, avoid visiting Dalit homes and are hesitant to handle infants or pregnant women because it requires close physical contact.(14)

The purpose of this study is to measure the factors associated with exclusive breastfeeding among Dalit mothers with children under the age of two. We anticipate that the findings of this research will serve as a valuable guideline for policymakers, stakeholders, and individuals in planning feasible interventions and strengthening existing factors related to exclusive breastfeeding. Ultimately, these effects aim to help Nepal achieve its national exclusive breastfeeding targets, thereby aligning with the broader aspirations of the Sustainable Development Goals for 2030. This research contributes to improving nutrition status (SDG2), prevents child mortality and lowers the risk of non-communicable disease (SDG3), and supports cognitive development and education (SDG4). Additionally, it underscores the role of breastfeeding in ending poverty, promoting economic growth, and reducing inequalities. Through a comprehensive exploration of exclusive breastfeeding within the context of Dalit communities, this study holds the potential to transform the lives of mothers and children, fostering a brighter and healthier future for all.

## Materials and Methods

### Study design

The cross-sectional analytical study design was used. Data was collected from 23^rd^ February 2023 to 8^th^ March 2023.

### Study Settings

The study was conducted in Dalit community of Rajbiraj Muicipality, Saptari District, Madhesh Province of Nepal.

A mid-sized municipality called Rajbiraj is found in the southern region of Nepal’s Madhesh Province. Rajbiraj is the district headquarters of Saptari and the eighth-largest city in the province. The total population of Rajbiraj Municipality is 68396, which is a larger population in comparison to other municipalities in the Saptari district. (15) In this district, there is an all-Dalit population. As a result, this setting was ideal for conducting research.

### Study Population and Sampling Frame

#### Study Population

The study population for this study was Dalit mothers of under 2 year children among Rajbiraj Municipality, Saptari, Nepal.

#### Sampling Frame

The list of mothers with children under 2 years was obtained from MIS the section of Rajbiraj Municipality.

### Sample Size

The Annual Report 2077/78 of Madhesh Province (DoHS<u>, 2078</u>) revealed that the overall prevalence of exclusive breastfeeding among children aged 0 to 6 months was 26%. While calculating the sample size, the allowable error was kept at 5% and the confidence level at 95%. The required sample size of the proposed study was calculated based on the aforesaid prevalence data.

Sample size was computed as

Sample size = (z^2^p*q/d^2^) (16)

Where,

Z= 1.96 reliability coefficient at alpha 0.05.

Prevalence (P) = 26% = 0.26 q = 1-P = 1-0.26 = 0.74

Allowable error (d) = 5% = 0.05

Applying the formula,

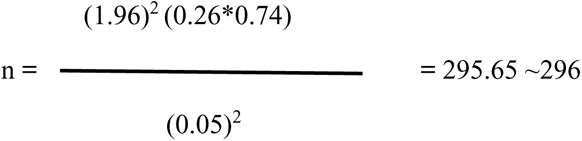

The required sample size for calculating the infinite population in calculated by considering the finite population,

Now, for a finite population,

Finite population (N) = 274 (which is the number of mothers of under 2 year children in Rajbiraj Municipality)

Sample size for finite population,

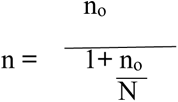

n = 296/1+296/274

= 142.30 ⁓142

Adding non – response rate of 10%, the required sample size = 156.2 ⁓ 156

### Sampling Techniques/Procedures

Simple random sampling technique was used to select the study participants. The sample frame for this study is 274 populations.

**Figure 3:**
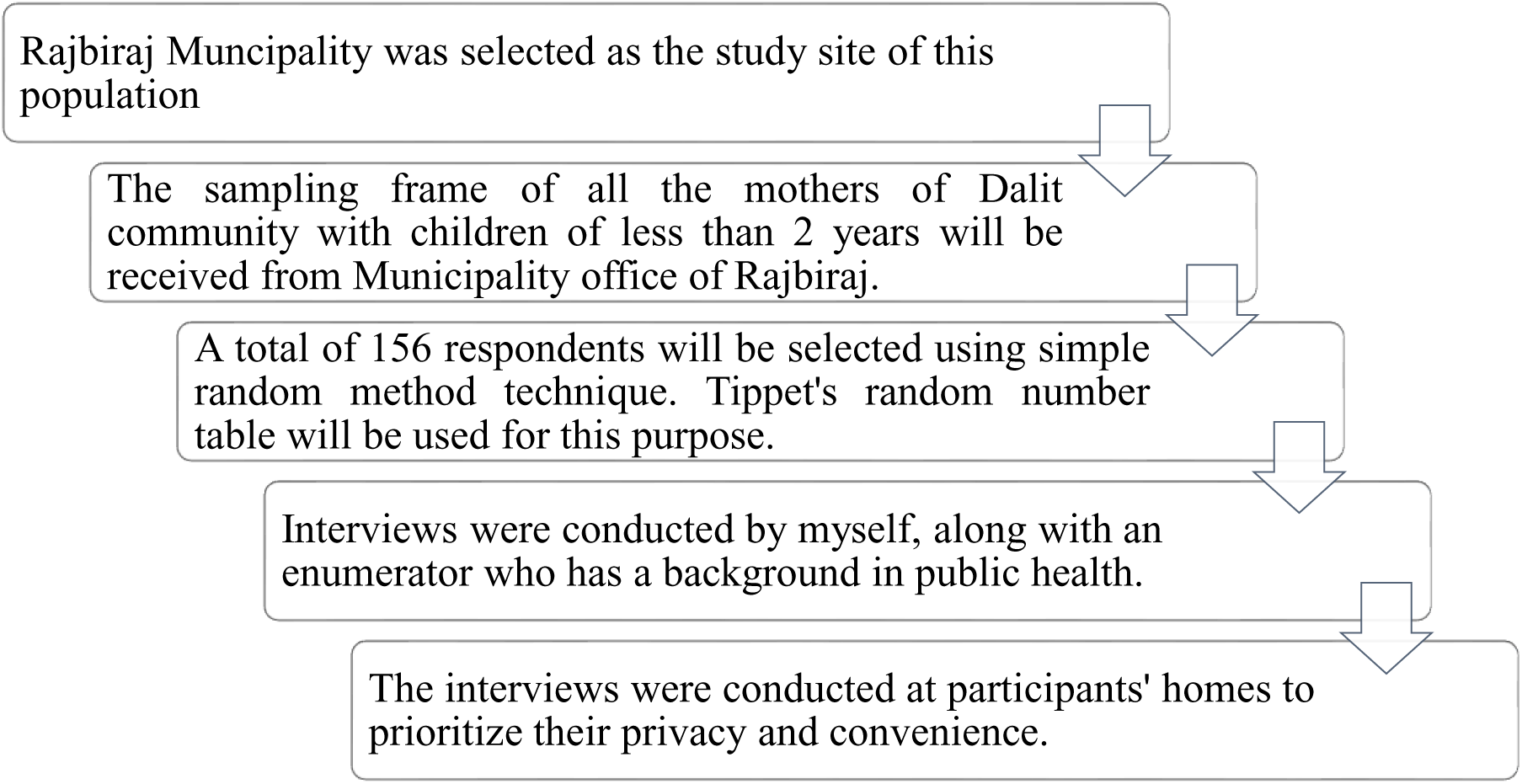
Sampling technique and procedure for the selection of the study participants.

### Data Collection Tools and Techniques

The data was collected using a semi-structured questionnaire, validated by a thorough review of the literature relevant to the study’s topic, and seeking the advice of a research expert. The research questionnaire was translated into Nepali and further translated into English. The questionnaire was divided into 2 parts. Information on sociodemographic characteristics was included in the first section, while exclusive breastfeeding was covered in the second. The feasibility of the questionnaire was maintained by pre-testing in the population of the sample. The actual data was gathered after the questionnaire has been improved. The face-to-face interview was conducted with mothers of under two years’ children in the Dalit community, Rajbiraj Municipality. Interviews were conducted by myself and an enumerator with a public health background. For illiterate participants, a witness was present during the consenting process, and the witness signed the informed consent form.

### Operational Definition

The following definitions were applied, consistent with WHO definitions:

#### Exclusive Breastfeeding

If an infant less than six months old took only breast milk and no additional food, water, or other liquids (with the exception of medicine and vitamins, if needed) continuing for the recommended 6 months.

#### Pre-lacteal feeding

Pre-lacteal feeding refers to the act of giving anything other than breast milk to an infant within the first three days after birth.

#### Dalit Community

Kalar, Kakaihiya, Kori, Khatik, Khatbe, Chamar, Chidimar, Dom, Tatma, Dusadh, Dhobi, Pattharkatta, Pasi, Bantar, Mushar, Mestar, and Sarvanga are Dalit castes of Madhesi origin who live in Rajbiraj municipality of Saptari District, Nepal. (3)

### Validity and Reliability of Tools

The content validity of the instrument was established by consultation with the study advisor’s subject experts, peers, and public health faculty research committee members at Nobel College. The questionnaire was modified as needed based on insightful opinions and ideas. Both English and Nepali was used in the design of the instrument. The original version of the tool was checked to ensure consistency in the meaning provided by the English and Nepali versions. After the interview is over, the completed questionnaire was carefully reviewed. A pretesting was conduction among 10% of the study sample before the main research to check feasibility and improve on the research tools. A pretesting was conduction among 16 randomly selected mothers of children under 2 years of age in Sambhunath municipality. Cronbach’s alpha coefficient was calculated selecting 16 questionnaires and used to measure consistency of tool. The Cronbach’s alpha of ® calculated was 0.806 which is greater than 0.7 and considered as standard and statistically acceptable.

### Data Management and Analysis

A data management plan was developed prior to data collection to make sure that the data are collected in the right format, are effectively organized, and are better interpreted. The day after the data is collected, the acquired data was cleaned, coded, and correctly entered in MS Excel. Data was manually verified, entered, and exported from MS Excel to IBM SPSS 20 for additional analysis. Analyses was performed for descriptive statistics like frequency, percentage, measures of central tendency, and measures of dispersion. Frequency tables was used for categorical variables, while mean and standard deviation was calculated for continuous variables. For the analysis involving testing for the association of independent variables with dependent variables, bivariate analysis (chi-square statistics and a p-value at <0.05) was used. Those that are significant in the bivariate analysis was further preceded by multivariate logistic regression. Independent variables with a P-value <0.05 was entered into the multivariate analysis.

### Ethical Consideration

Before collecting data, administrative approval was obtained from the concerned authority at Nobel College. Then, formal approval and ethical clearance was obtained from the Institutional Review Committee of Nobel College. Permission was obtained from the Rajbiraj Municipality Office for conducting the purposed study. Written Informed consent was obtained from the participants prior to data collection, assuring voluntary participation and that the individual’s identity was not disclosed in the report. Confidentiality was maintained throughout the study period, and the collected data was used only for research purposes.

## Results/Findings

This chapter deals with the result tabulation and interpretation of the data obtained from 156 Dalit mothers of under 2 years’ children who voluntarily participated in his study.

### Univariate Analysis

The study, encompassing 156 respondents, revealed that the mothers’ ages ranged from 19 to 36 years, with a mean age of 24.59 years (confidence interval: 24.03 to 25.15). Over 60% fell within the 19 to 24-year age group. Children’s ages ranged from 6 to 24 months, with a mean age of 11.39 months (confidence interval: 10.64 to 12.14). The majority of respondents (64.1%) had children in the 6 to 12-month age group. In terms of gender, 59.6% were male. Most respondents were homemakers (84%), with husbands predominantly working as laborers (57.7%). Most of mothers had completed secondary education (48.1%) while Husbands’ education receiving secondary education (55.1%). Nearly 59% belonged to nuclear families, and the majority had two children (41.7%). Lifestyle-wise, 71.8% didn’t smoke, and 85.9% abstained from alcohol.

Table 4 illustrates the distribution of the population’s knowledge on breastfeeding. Among 156 respondents 57.1% had sufficient knowledge about breastfeeding, while 42.9% had no knowledge. Of the 89 respondents with knowledge, 78.7% were aware of the benefits, 73% knew about exclusive breastfeeding, but only 19% were informed about managing breast-related problems. Health workers were the primary source of information (83.1%), followed by friends and community (24.7%), social media (29.2%), and mothers’ group meetings (29.2%).

**Table 3:**
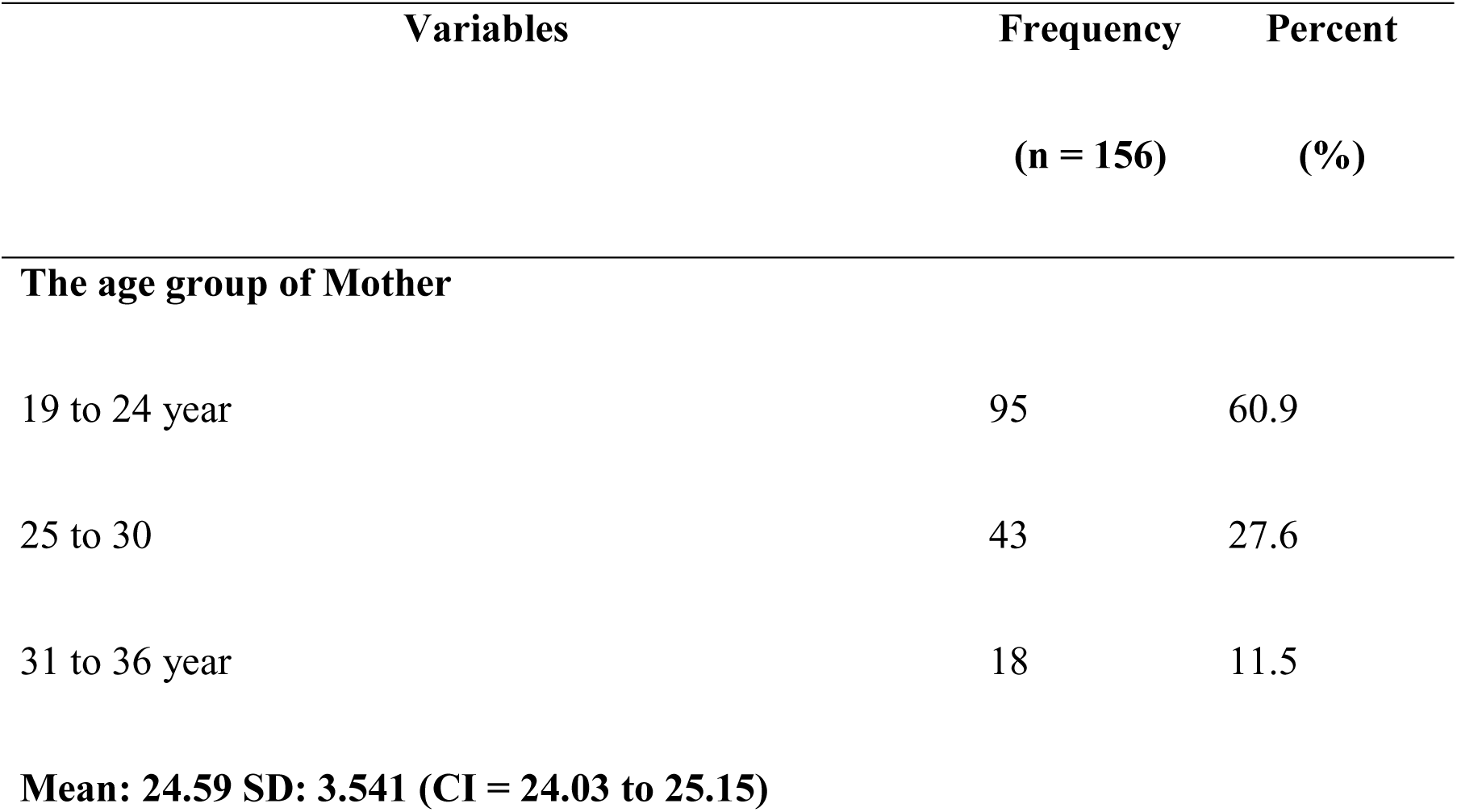
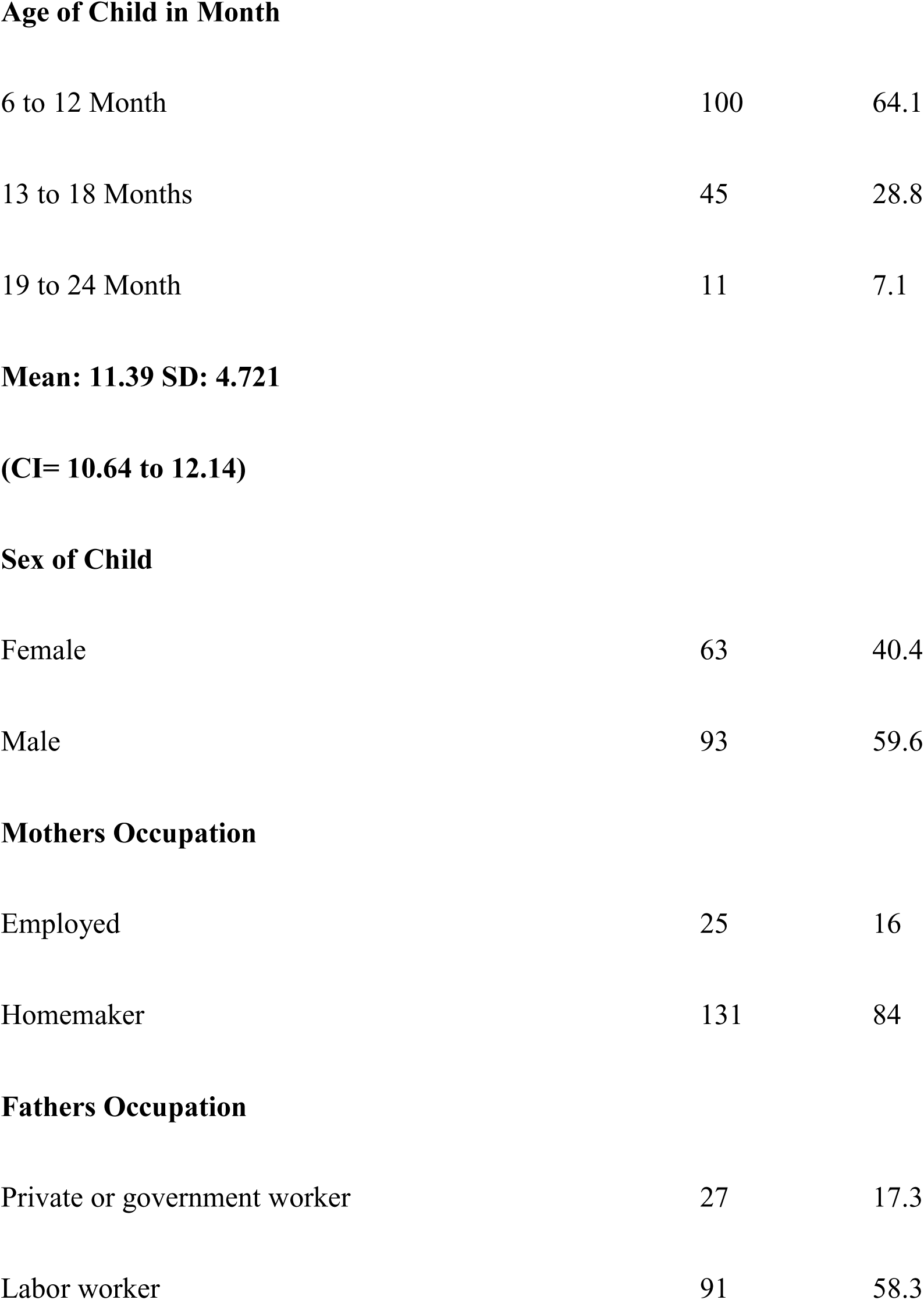
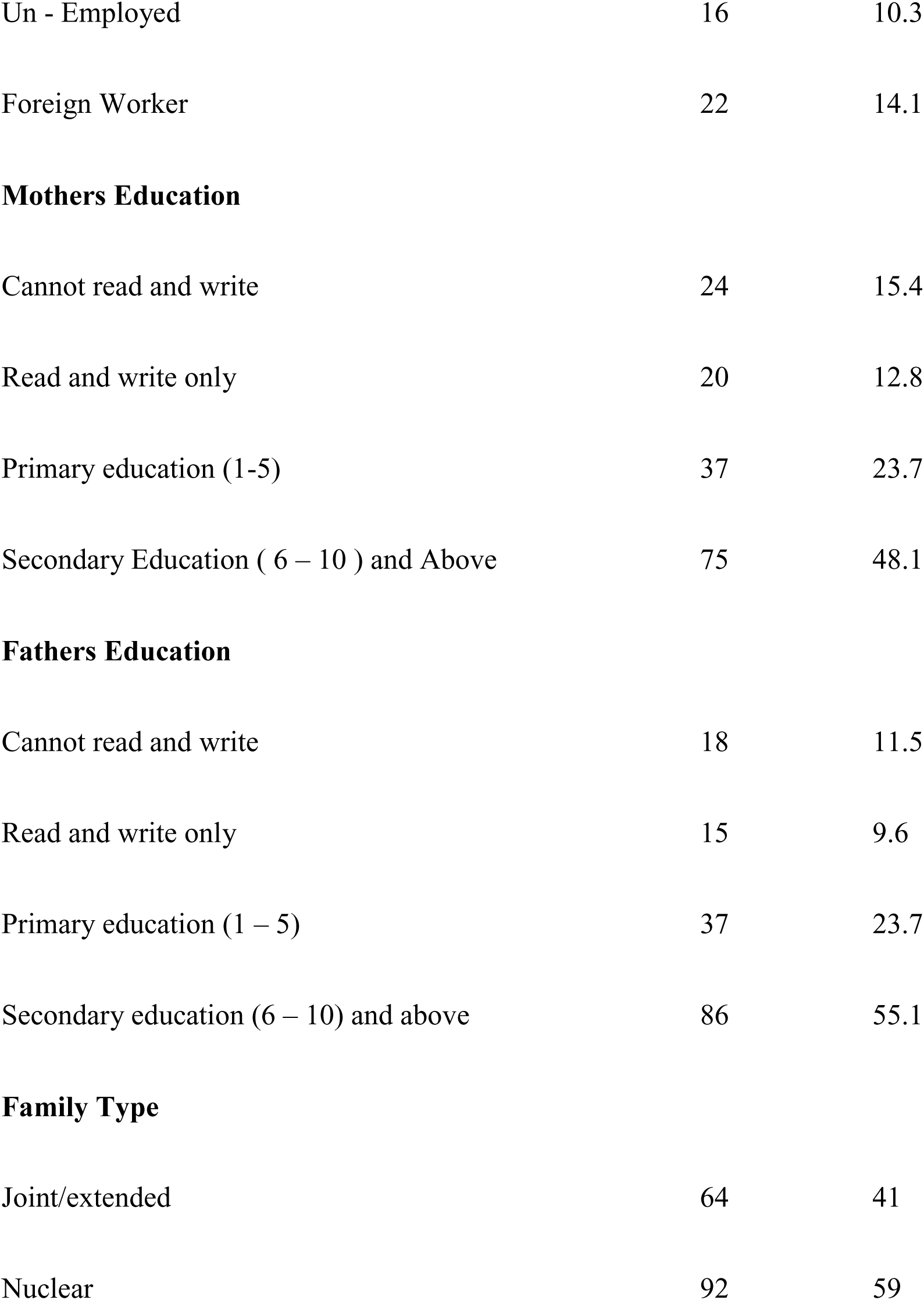
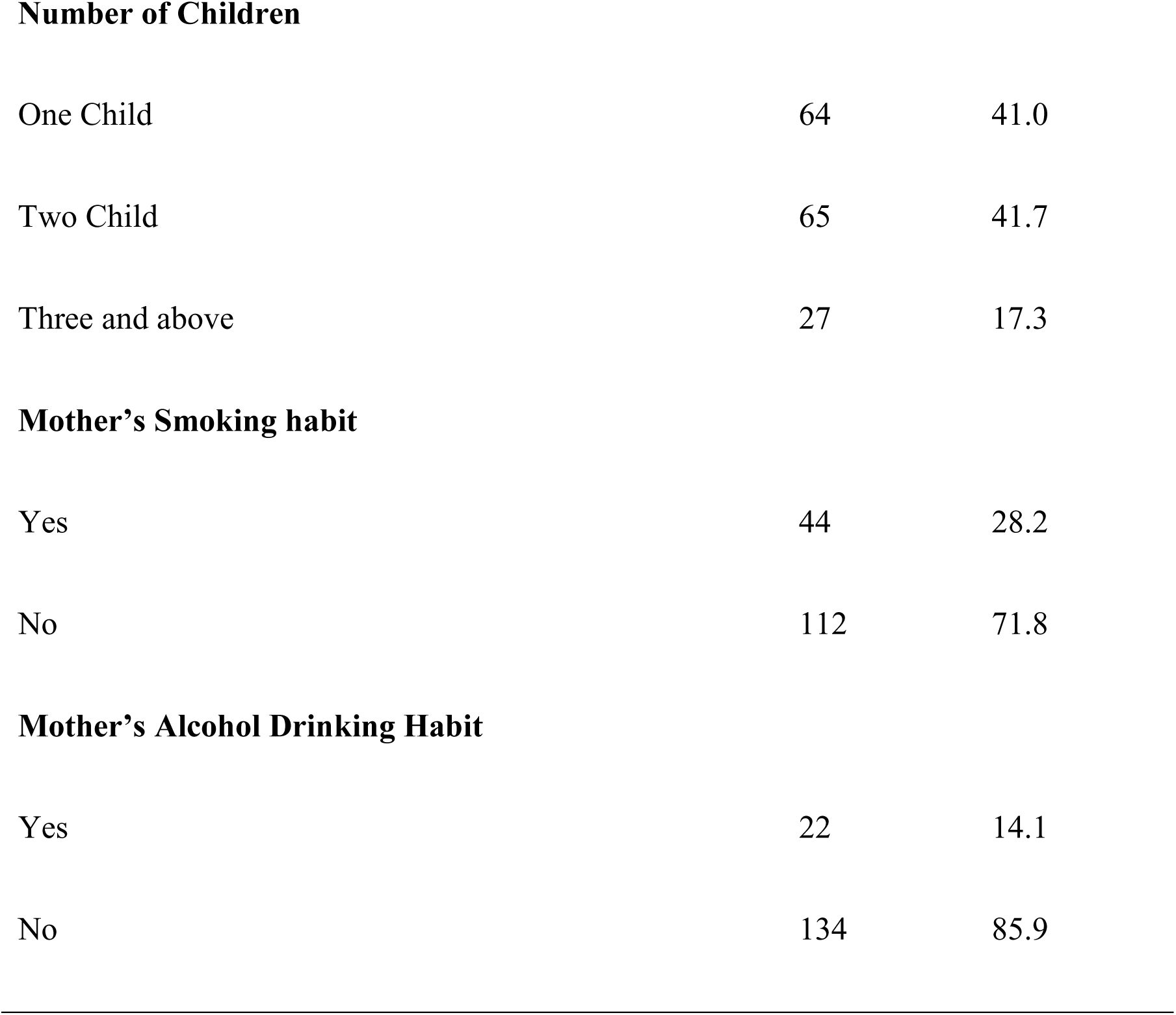
Sociodemographic variable (n=156)

| **Variables** | **Frequency** | **Percent** |
| --- | --- | --- |
|  | **(n = 156)** | **(%)** |
| **The age group of Mother** |  |  |
| 19 to 24 year | 95 | 60.9 |
| 25 to 30 | 43 | 27.6 |
| 31 to 36 year | 18 | 11.5 |
| **Age of Child in Month** |  |  |
| 6 to 12 Month | 100 | 64.1 |
| 13 to 18 Months | 45 | 28.8 |
| 19 to 24 Month | 11 | 7.1 |
| **Sex of Child** |  |  |
| Female | 63 | 40.4 |
| Male | 93 | 59.6 |
| **Mothers Occupation** |  |  |
| Employed | 25 | 16 |
| Homemaker | 131 | 84 |
| **Fathers Occupation** |  |  |
| Private or government worker | 27 | 17.3 |
| Labor worker | 91 | 58.3 |
| Un - Employed | 16 | 10.3 |
| Foreign Worker | 22 | 14.1 |
| **Mothers Education** |  |  |
| Cannot read and write | 24 | 15.4 |
| Read and write only | 20 | 12.8 |
| Primary education (1-5) | 37 | 23.7 |
| Secondary Education ( 6 – 10 ) and Above | 75 | 48.1 |
| **Fathers Education** |  |  |
| Cannot read and write | 18 | 11.5 |
| Read and write only | 15 | 9.6 |
| Primary education (1 – 5) | 37 | 23.7 |
| Secondary education (6 – 10) and above | 86 | 55.1 |
| **Family Type** |  |  |
| Joint/extended | 64 | 41 |
| Nuclear | 92 | 59 |
| **Number of Children** |  |  |
| One Child | 64 | 41.0 |
| Two Child | 65 | 41.7 |
| Three and above | 27 | 17.3 |
| **Mother's Smoking habit** |  |  |
| Yes | 44 | 28.2 |
| No | 112 | 71.8 |
| **Mother's Alcohol Drinking Habit** |  |  |
| Yes | 22 | 14.1 |
| No | 134 | 85.9 |

**Table 4:** Knowledge about Exclusive breastfeeding.

| **Variables** | **Frequency** | **Percent** |
| --- | --- | --- |
|  | **(n = 156)** | **(%)** |
| **Knowledge about Breastfeeding** |  |  |
| Yes | 89 | 57.1 |
| No | 67 | 42.9 |
| **(n=89)** |  |  |
| **Knowledge related information** |  |  |
| Benefits of Breastfeeding | 70 | 78.7 |
| Exclusive Breastfeeding | 65 | 73 |
| Management of breast-related problems | 19 | 19 |
| **Source of information** |  |  |
| Health worker | 74 | 83.1 |
| Friends and Community | 22 | 24.7 |
| Social Media | 26 | 29.2 |
| Meeting of Mothers' group | 26 | 29.2 |

Table 5 outlines the feeding practices observed among infants in the study, revealing that a significant majority (68.6%) were breastfed within one hour of birth, indicating a strong adherence to early breastfeeding initiation. Concerning pre-lacteal feeding, over four-fifths (82.7%) did not receive any pre-lacteal feed during their first three days of life. As for colostrum milk, nearly four-fifths (79.5%) were fed colostrum, while 20.5% were not. Regarding the introduction of solid or liquid food, 20.5% introduced them to their children between 0 to 2 months, 35.9% between 3 to 5 months, and the remaining 43.6% introduced complementary feeding at 6 months or later.

**Table 5:**
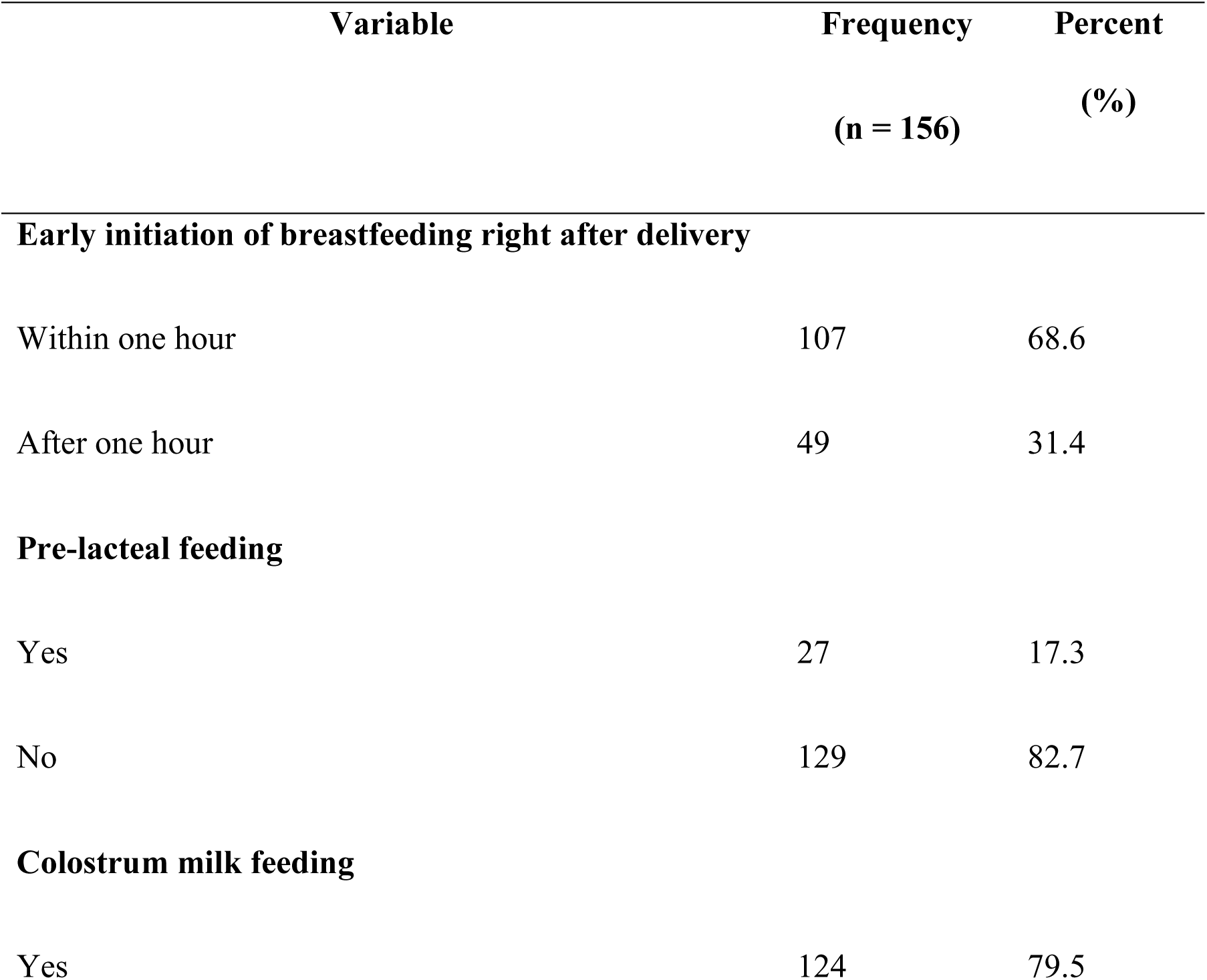
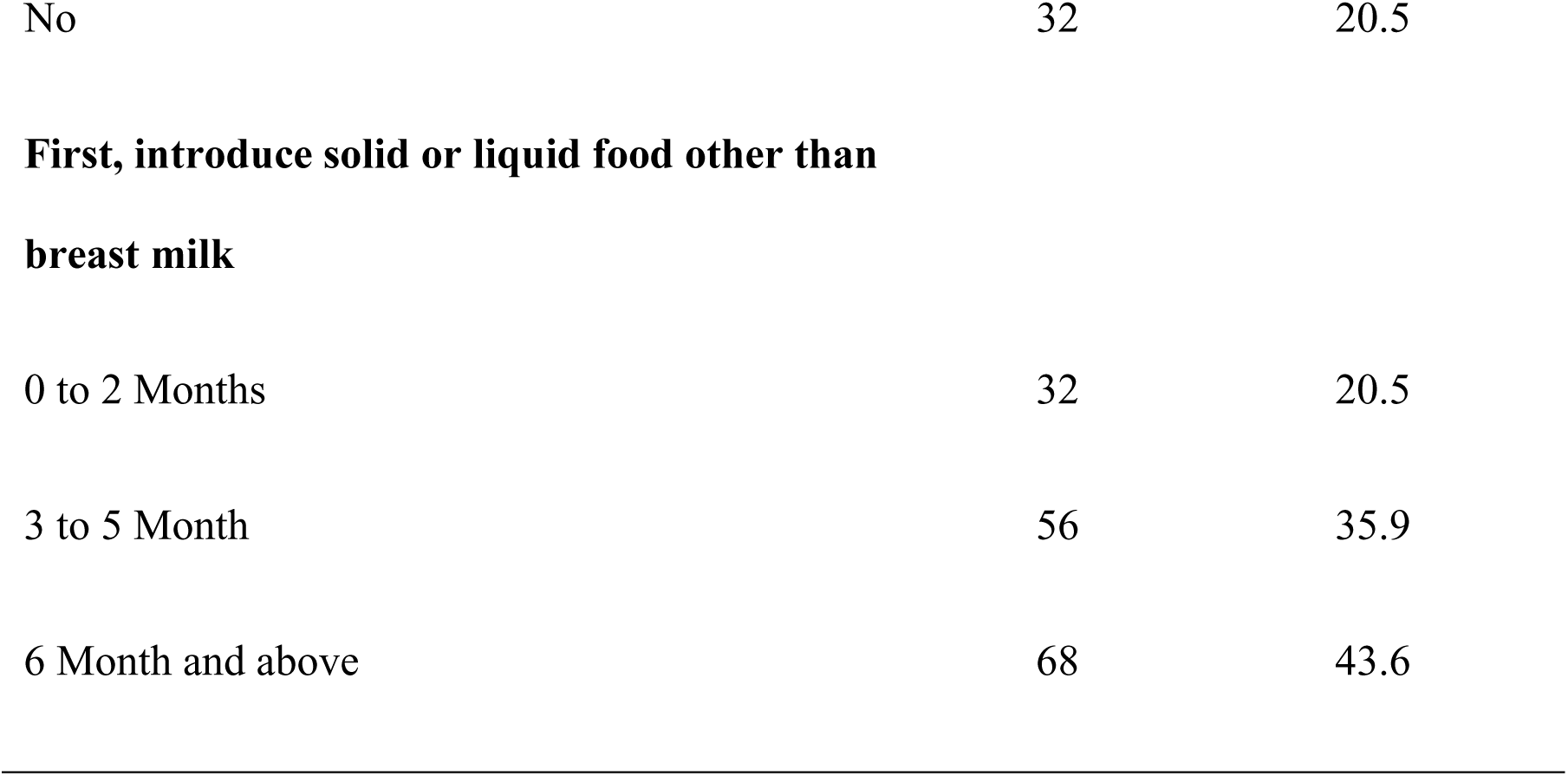
Child-related information.

| Variable | Frequency<br>(n = 156) | Percent<br>(%) |
| --- | --- | --- |
| **Early initiation of breastfeeding right after delivery** |  |  |
| Within one hour | 107 | 68.6 |
| After one hour | 49 | 31.4 |
| **Pre-lacteal feeding** |  |  |
| Yes | 27 | 17.3 |
| No | 129 | 82.7 |
| **Colostrum milk feeding** |  |  |
| Yes | 124 | 79.5 |
| No | 32 | 20.5 |
| **First, introduce solid or liquid food other than breast milk** |  |  |
| 0 to 2 Months | 32 | 20.5 |
| 3 to 5 Month | 56 | 35.9 |
| 6 Month and above | 68 | 43.6 |

Table 6 displays mother-related information linked to exclusive breastfeeding, indicating that all 156 respondents attended health centers for ANC, with 59.6% visiting four or more times. Concerning postnatal care (PNC), 66.7% did not receive PNC services, and 33.3% did. About 47.4% received exclusive breastfeeding counseling during ANC and PNC visits, while 52.6% did not. For childbirth practices, 92.9% delivered at a health facility, predominantly through spontaneous vaginal delivery (87.2%), with 12.8% undergoing a cesarean section.

**Table 6:**
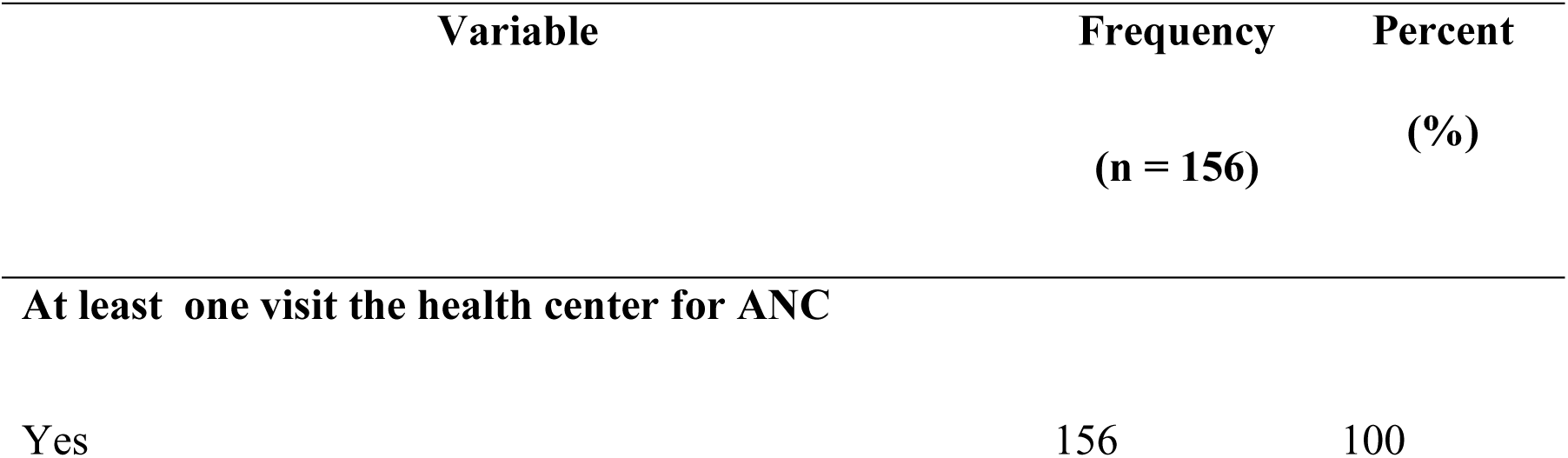
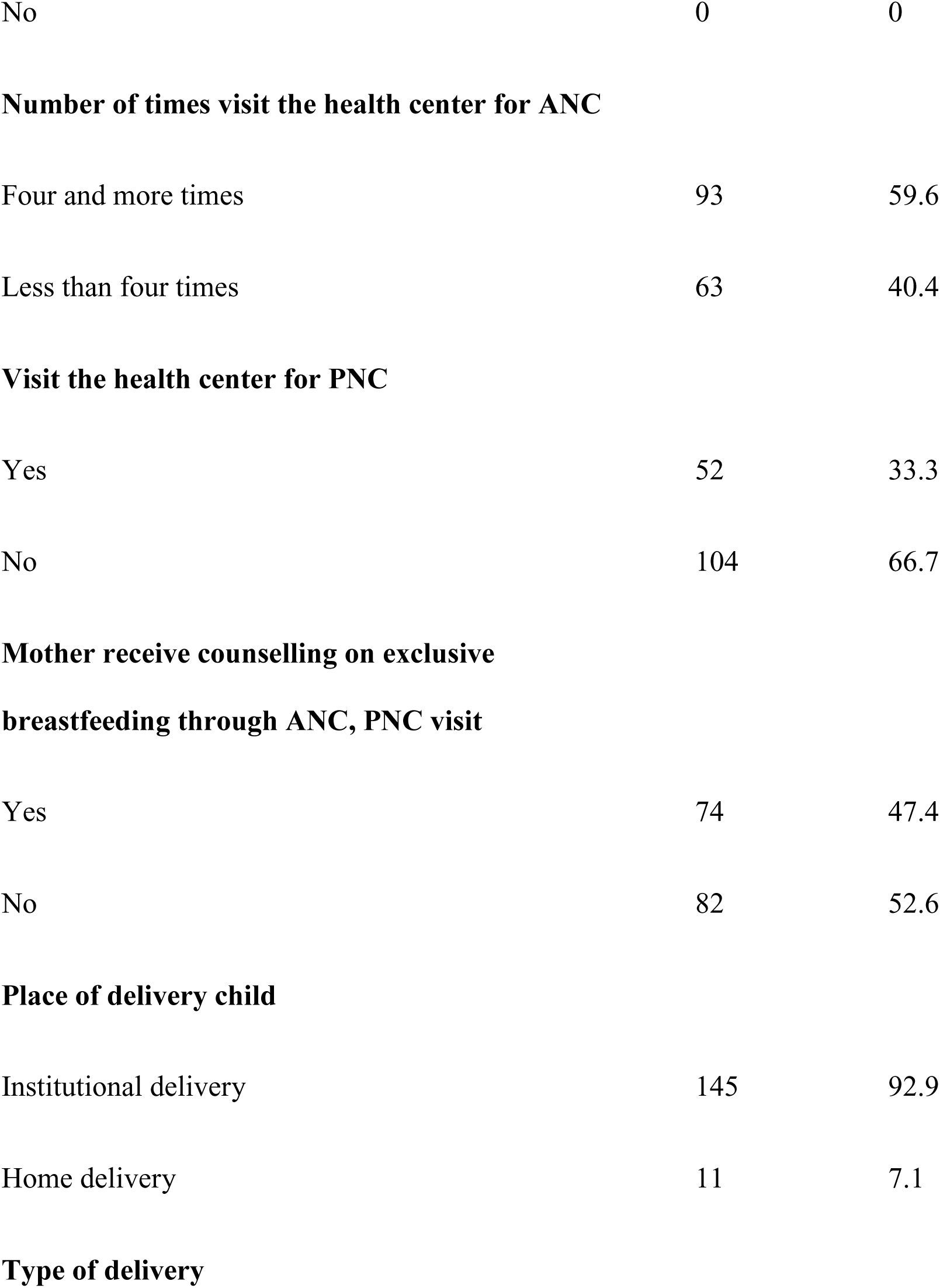
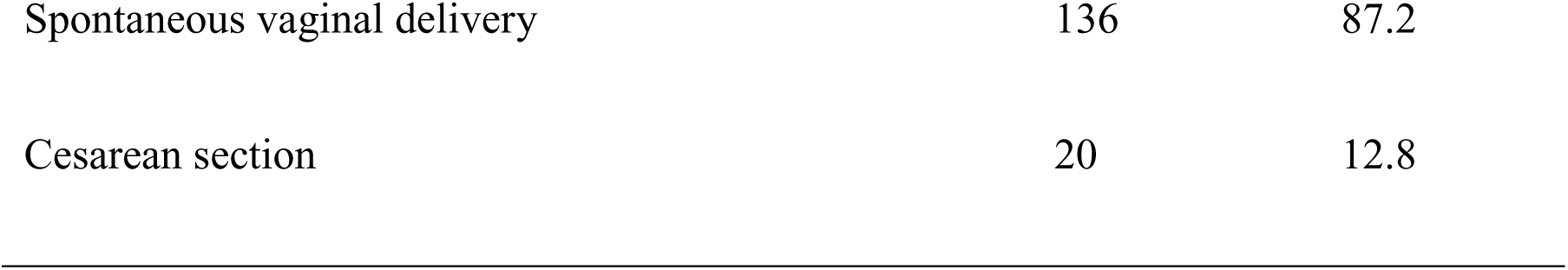
Mother-related information.

| Variable | Frequency<br>(n = 156) | Percent<br>(%) |
| --- | --- | --- |
| **At least one visit the health center for ANC** |  |  |
| Yes | 156 | 100 |
| No | 0 | 0 |
| **Number of times visit the health center for ANC** |  |  |
| Four and more times | 93 | 59.6 |
| Less than four times | 63 | 40.4 |
| **Visit the health center for PNC** |  |  |
| Yes | 52 | 33.3 |
| No | 104 | 66.7 |
| **Mother receive counselling on exclusive**<br>**breastfeeding through ANC, PNC visit** |  |  |
| Yes | 74 | 47.4 |
| No | 82 | 52.6 |
| **Place of delivery child** |  |  |
| Institutional delivery | 145 | 92.9 |
| Home delivery | 11 | 7.1 |
| **Type of delivery** |  |  |
| Spontaneous vaginal delivery | 136 | 87.2 |
| Cesarean section | 20 | 12.8 |

In Table 7, the majority (56.4%) of respondents did not practice exclusive breastfeeding, while 43.6% reported adhering to exclusive breastfeeding. Of the 68 respondents practicing exclusive breastfeeding, 42.6% were influenced by health workers, 35.3% made the decision independently, and 22.1% were influenced by family, friends, or the community.

**Table 7:**
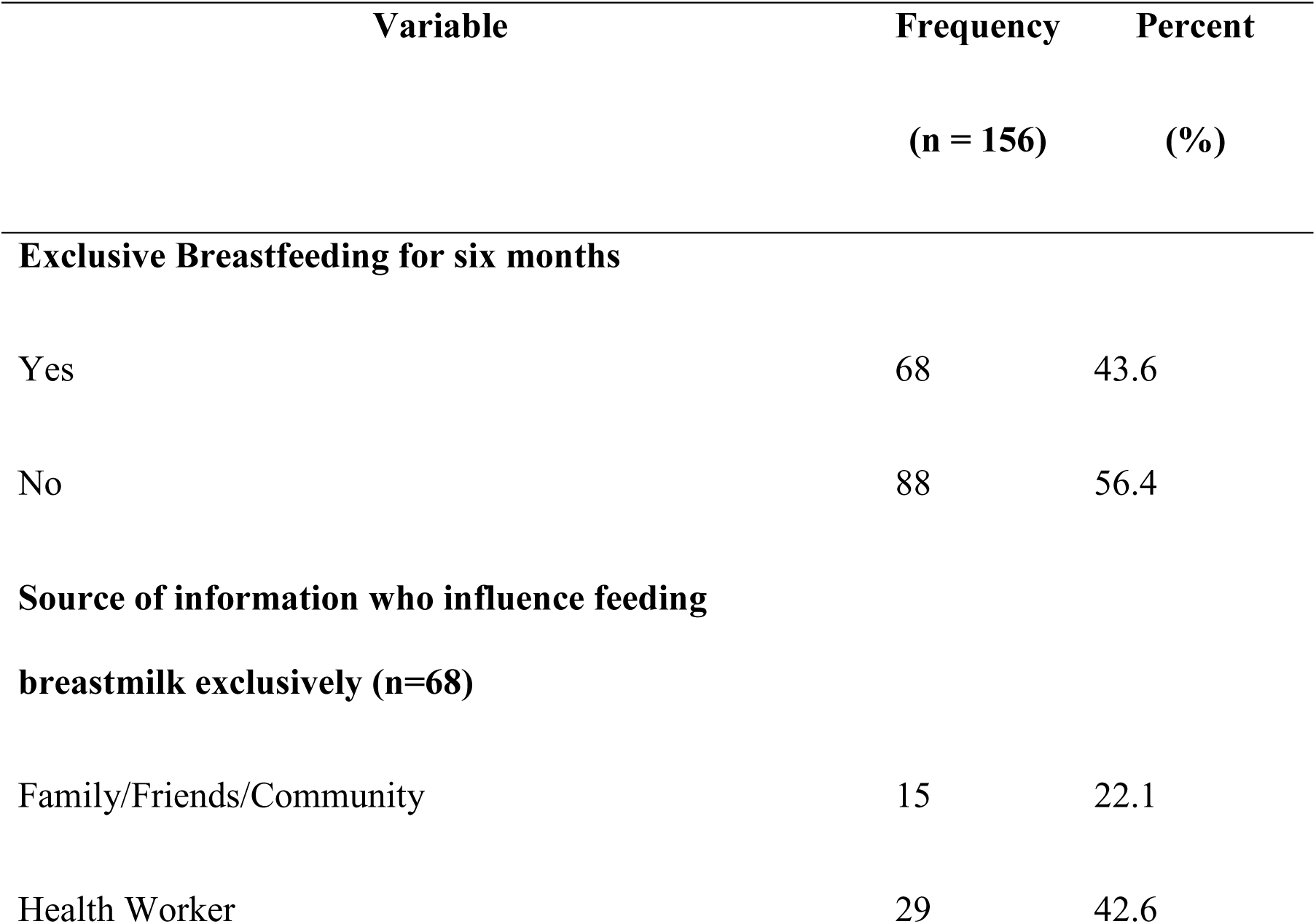
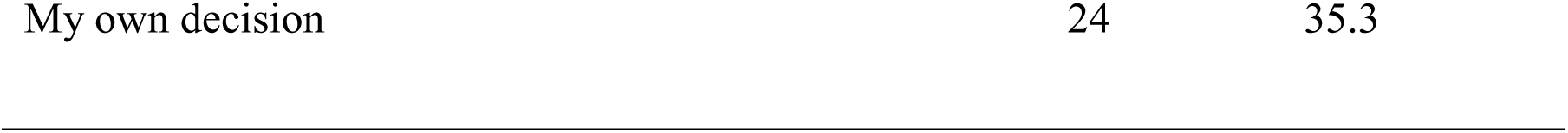
Exclusive breastfeeding-related information.

| Variable | Frequency<br>(n = 156) | Percent<br>(%) |
| --- | --- | --- |
| **Exclusive Breastfeeding for six months** |  |  |
| Yes | 68 | 43.6 |
| No | 88 | 56.4 |
| **Source of information who influence feeding**<br>**breastmilk exclusively (n=68)** |  |  |
| Family/Friends/Community | 15 | 22.1 |
| Health Worker | 29 | 42.6 |
| My own decision | 24 | 35.3 |

**Table 8:** Factors to discontinue exclusive breastfeeding.

| Variable | Frequency<br>(n=88) | Percent<br>(%) |
| --- | --- | --- |
| I had trouble getting the milk flow to start | 59 | 67 |
| I didn't have enough milk | 54 | 61.4 |
| Breast milk does not satisfy my baby | 52 | 59.1 |
| I had too many household duties | 30 | 34.1 |
| I thought my baby was not gaining enough weight | 26 | 29.5 |
| My baby had trouble sucking or latching | 16 | 18.8 |
| My baby become sick and could not breastfeed | 12 | 13.6 |

Among the total respondents (88) not practicing exclusive breastfeeding, reasons for discontinuation were documented in Table 6. The primary reason, cited by 67% of respondents, was difficulty initiating milk flow. Additionally, 61.4% reported insufficient milk production by the mother, while 59.1% felt breast milk did not satisfy the baby. Other factors included excessive household duties (34.1%), maternal concerns about inadequate weight gain (29.5%), difficulties with the baby’s sucking or latching (18.8%), and the baby falling ill and being unable to breastfeed (13.6%).

### Bivariate Analysis

Table 9 presents data on exclusive breastfeeding among 156 respondents, with 43.6% practicing it and 56.4% not. Within the exclusive breastfeeding group, age distribution shows 52.9% in the 19 to 24-year range, 32.4% in the 25 to 30-year range, and 14.7% in the 31 to 36-year range. No significant association with age was found (P = 0.079). Sex of the child revealed a significant association (P = 0.033), with 69.1% of exclusively breastfed being male. Mothers’ occupation also showed significance (P = 0.031), with 91.2% of homemakers exclusively breastfeeding. Mother Education level exhibited a significant association (P = 0.049), with 36.7% of those completing secondary education and above practicing exclusive breastfeeding. Family type and smoking habits also revealed significant associations (P = 0.024 and P = 0.027, respectively), while alcohol-drinking habits did not show an association (P = 0.230).

**Table 9:**
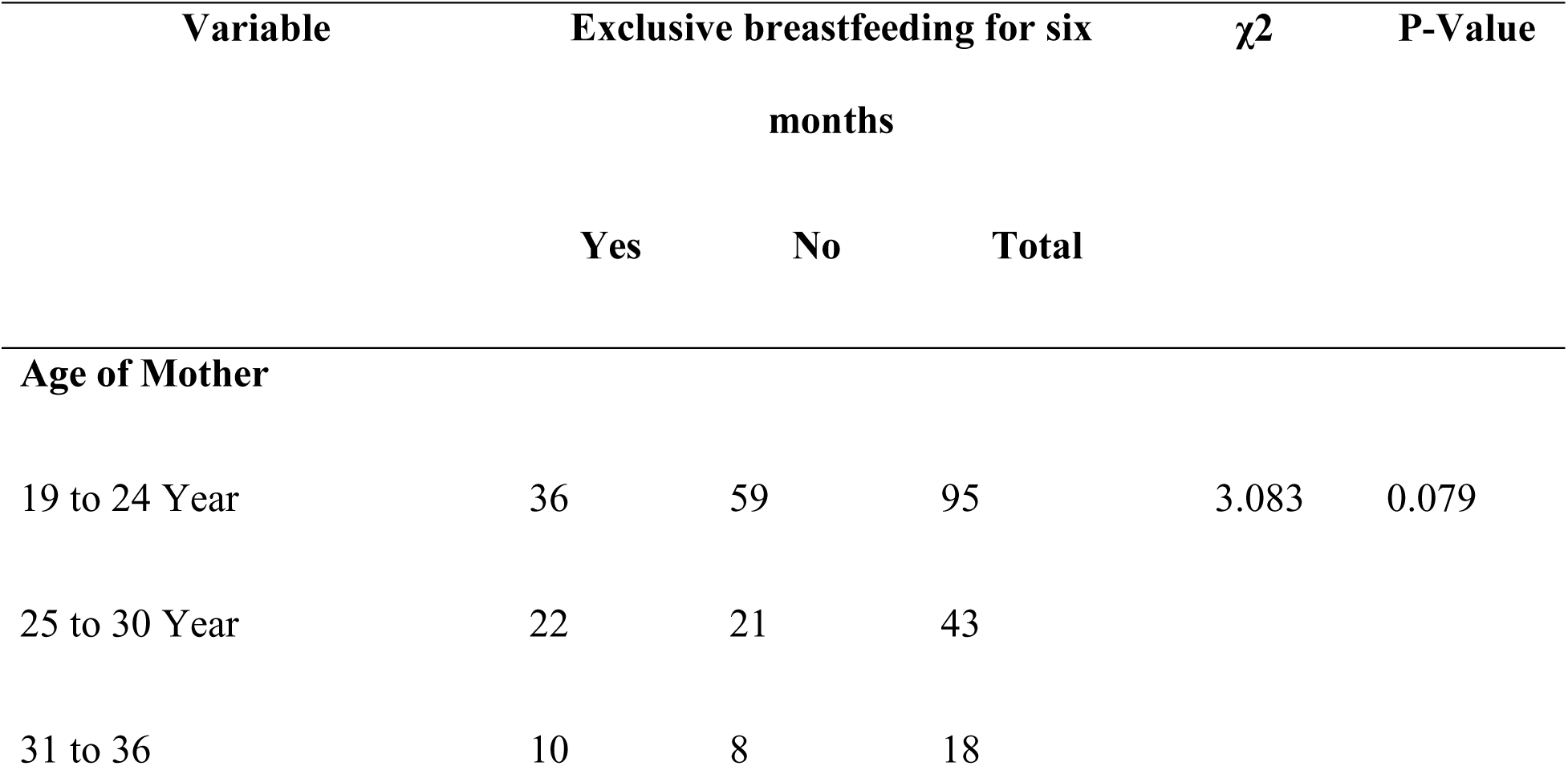
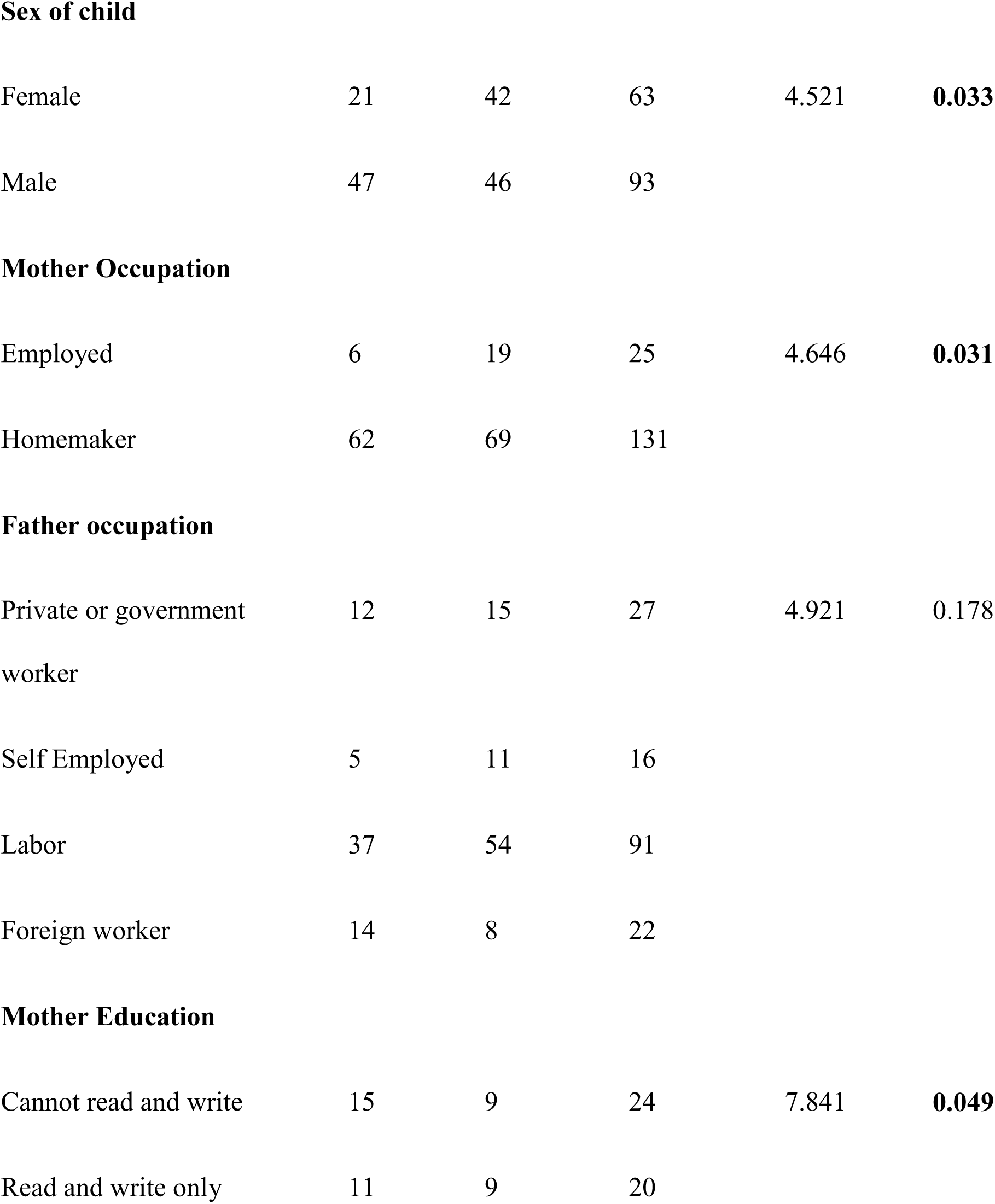
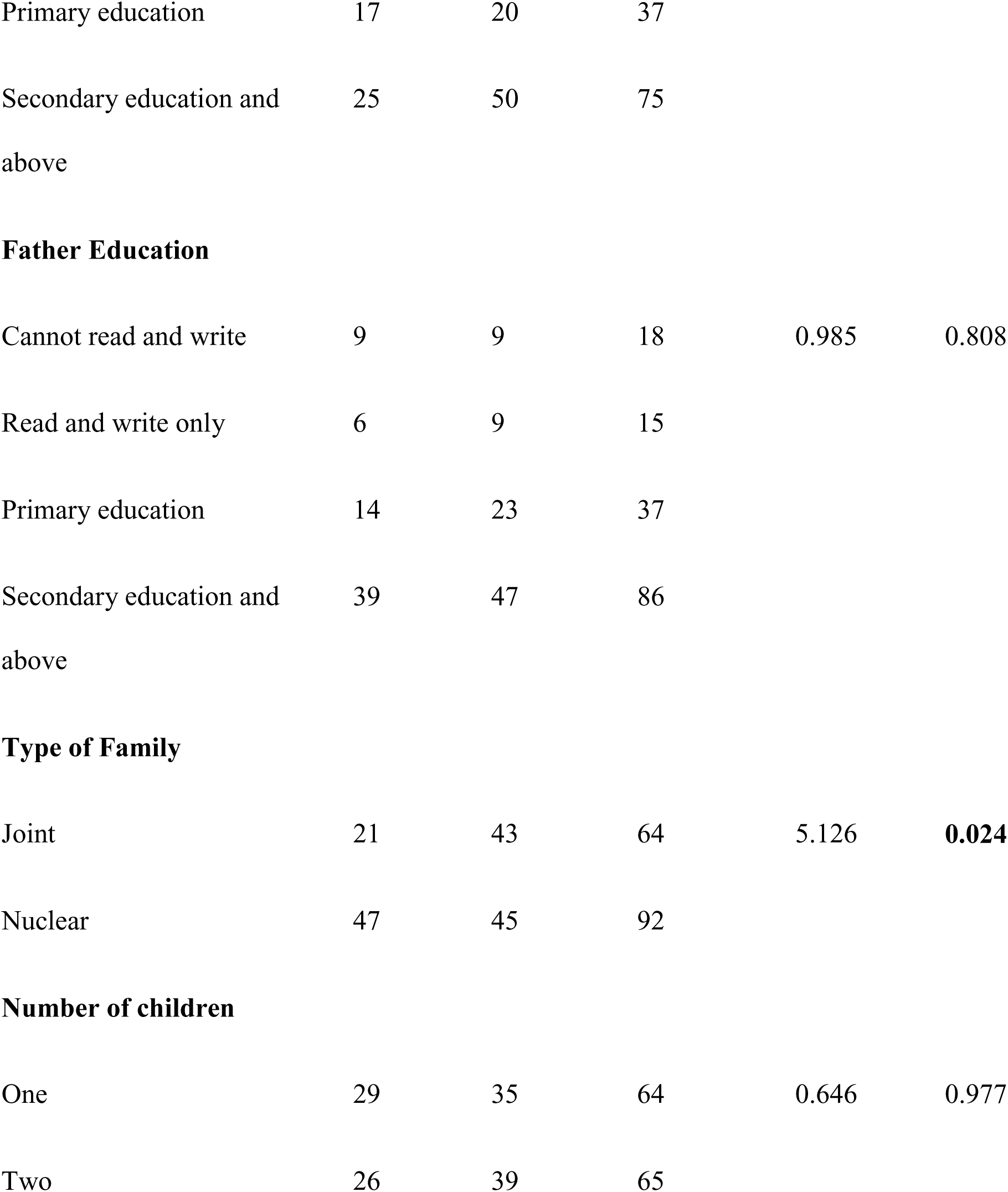
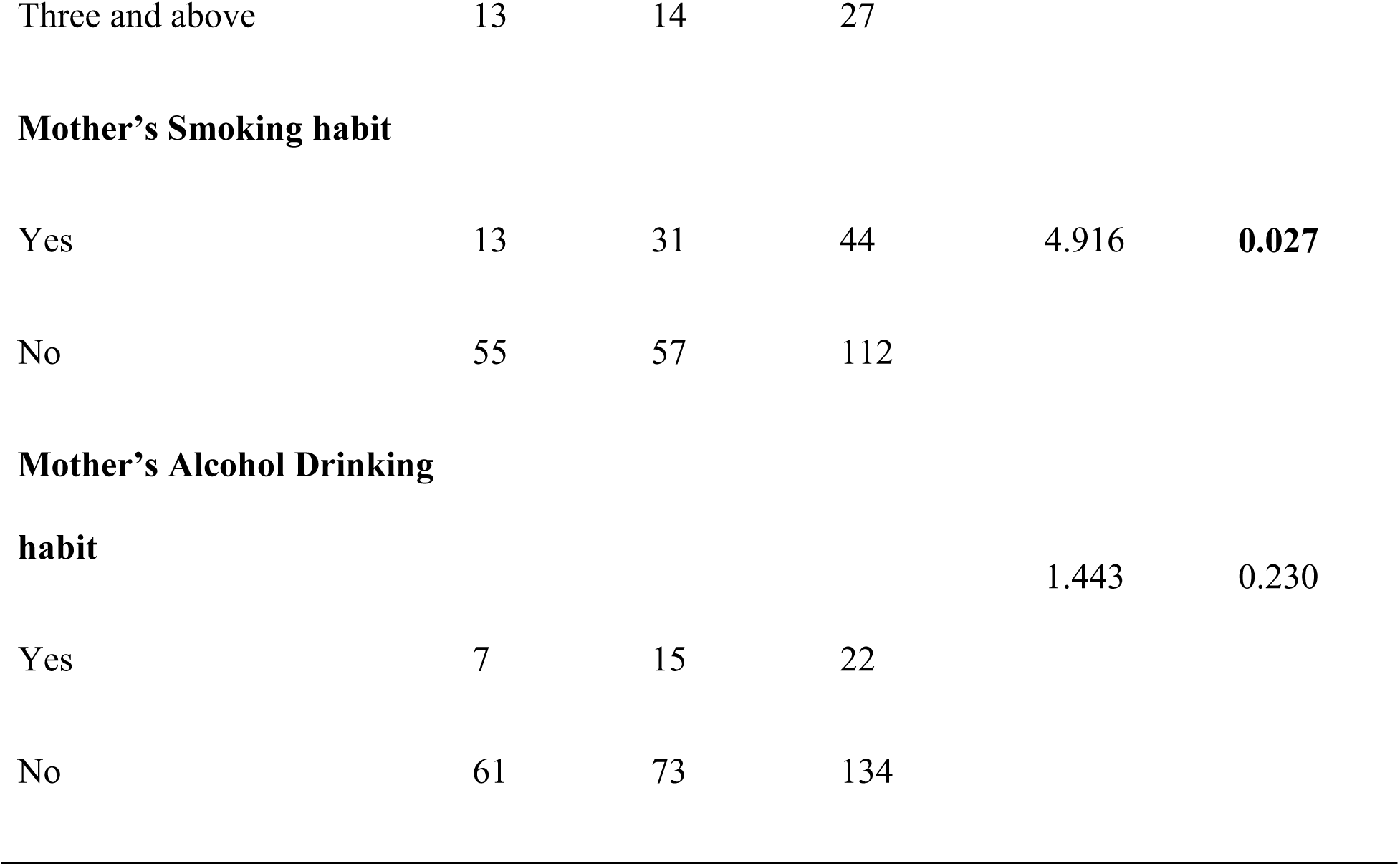
Association between exclusive breastfeeding and selected sociodemographic variables.

| Variable | Exclusive breastfeeding for six months | | | $\chi^2$ | P-Value |
| --- | --- | --- | --- | --- | --- |
|  | Yes | No | Total |  |  |
| Age of Mother |  |  |  |  |  |
| 19 to 24 Year | 36 | 59 | 95 | 3.083 | 0.079 |
| 25 to 30 Year | 22 | 21 | 43 |  |  |
| 31 to 36 | 10 | 8 | 18 |  |  |
| **Sex of child** |  |  |  |  |  |
| Female | 21 | 42 | 63 | 4.521 | **0.033** |
| Male | 47 | 46 | 93 |  |  |
| **Mother Occupation** |  |  |  |  |  |
| Employed | 6 | 19 | 25 | 4.646 | **0.031** |
| Homemaker | 62 | 69 | 131 |  |  |
| **Father occupation** |  |  |  |  |  |
| Private or government<br>worker | 12 | 15 | 27 | 4.921 | 0.178 |
| Self Employed | 5 | 11 | 16 |  |  |
| Labor | 37 | 54 | 91 |  |  |
| Foreign worker | 14 | 8 | 22 |  |  |
| **Mother Education** |  |  |  |  |  |
| Cannot read and write | 15 | 9 | 24 | 7.841 | **0.049** |
| Read and write only | 11 | 9 | 20 |  |  |

|  |  |  |  |
| --- | --- | --- | --- |
| Primary education | 17 | 20 | 37 |
| Secondary education and<br>above | 25 | 50 | 75 |
| **Father Education** |  |  |  |  |  |
| Cannot read and write | 9 | 9 | 18 | 0.985 | 0.808 |
| Read and write only | 6 | 9 | 15 |  |  |
| Primary education | 14 | 23 | 37 |  |  |
| Secondary education and<br>above | 39 | 47 | 86 |  |  |
| **Type of Family** |  |  |  |  |  |
| Joint | 21 | 43 | 64 | 5.126 | **0.024** |
| Nuclear | 47 | 45 | 92 |  |  |
| **Number of children** |  |  |  |  |  |
| One | 29 | 35 | 64 | 0.646 | 0.977 |
| Two | 26 | 39 | 65 |  |  |
| Three and above | 13 | 14 | 27 |  |  |
| **Mother's Smoking habit** |  |  |  |  |  |
| Yes | 13 | 31 | 44 | 4.916 | **0.027** |
| No | 55 | 57 | 112 |  |  |
| **Mother's Alcohol Drinking habit** |  |  |  |  |  |
|  |  |  |  | 1.443 | 0.230 |
| Yes | 7 | 15 | 22 |  |  |
| No | 61 | 73 | 134 |  |  |

Table 10 examines the association between exclusive breastfeeding and various factors among 156 respondents. Of the 68 (43.6%) exclusively breastfeeding respondents, 69.1% had knowledge about breastfeeding, showing a significant association (P = 0.007). Among those with knowledge about breastfeeding, exclusive breastfeeding was significantly associated with knowledge about the benefits of breastfeeding (P = 0.04) and knowledge about exclusive breastfeeding (P = 0.007). The study found a significant association between exclusive breastfeeding and avoiding pre-lacteal feeding (P = 0.0001) and practicing colostrum milk feeding (P = 0.0001). Additionally, the number of ANC visits was significantly associated with exclusive breastfeeding (P = 0.033), with 60.3% of exclusively breastfeeding mothers receiving counseling on the topic (P = 0.005). Although no significant association was found, the type of delivery showed a trend towards significance (P = 0.073).

**Table 10:**
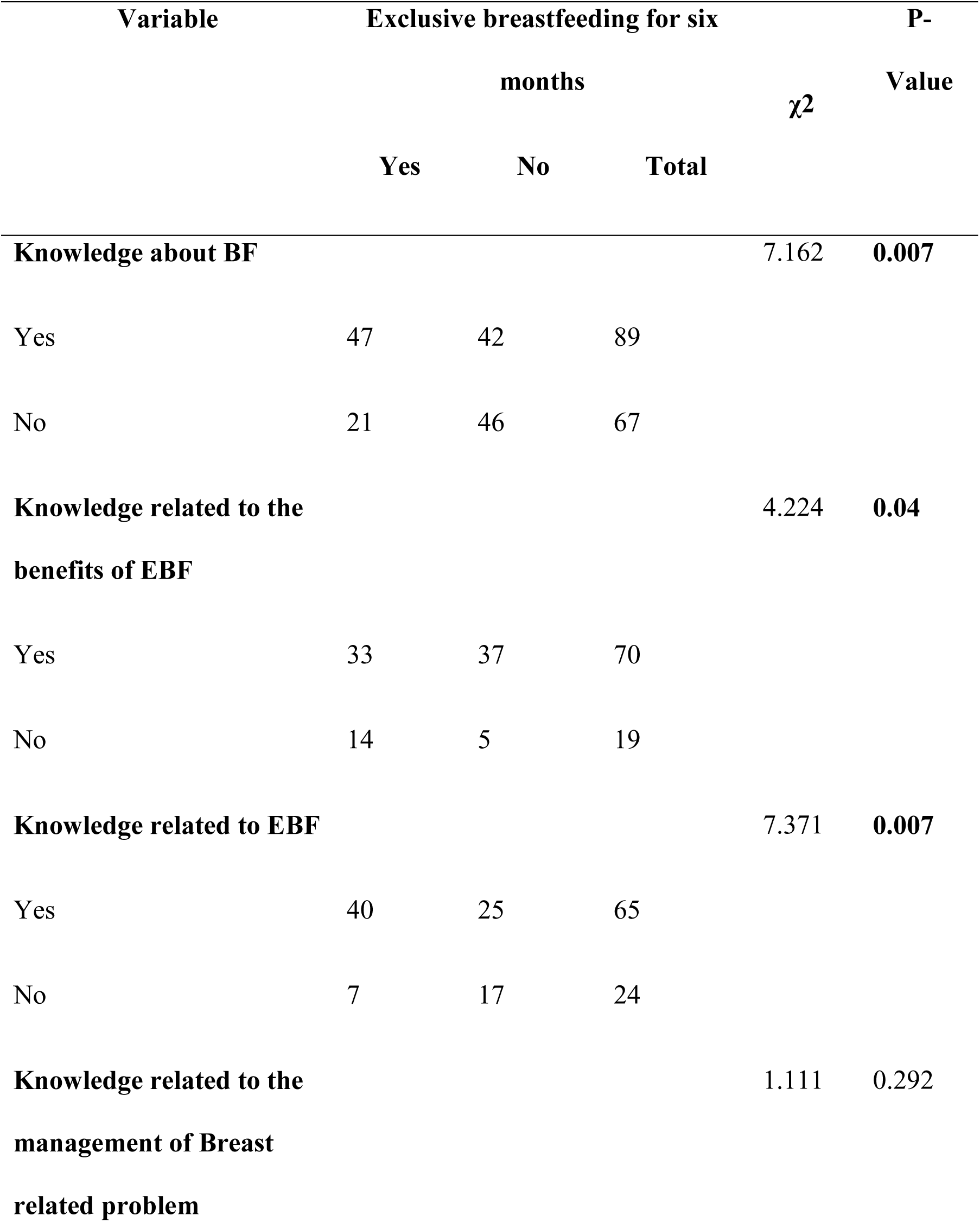
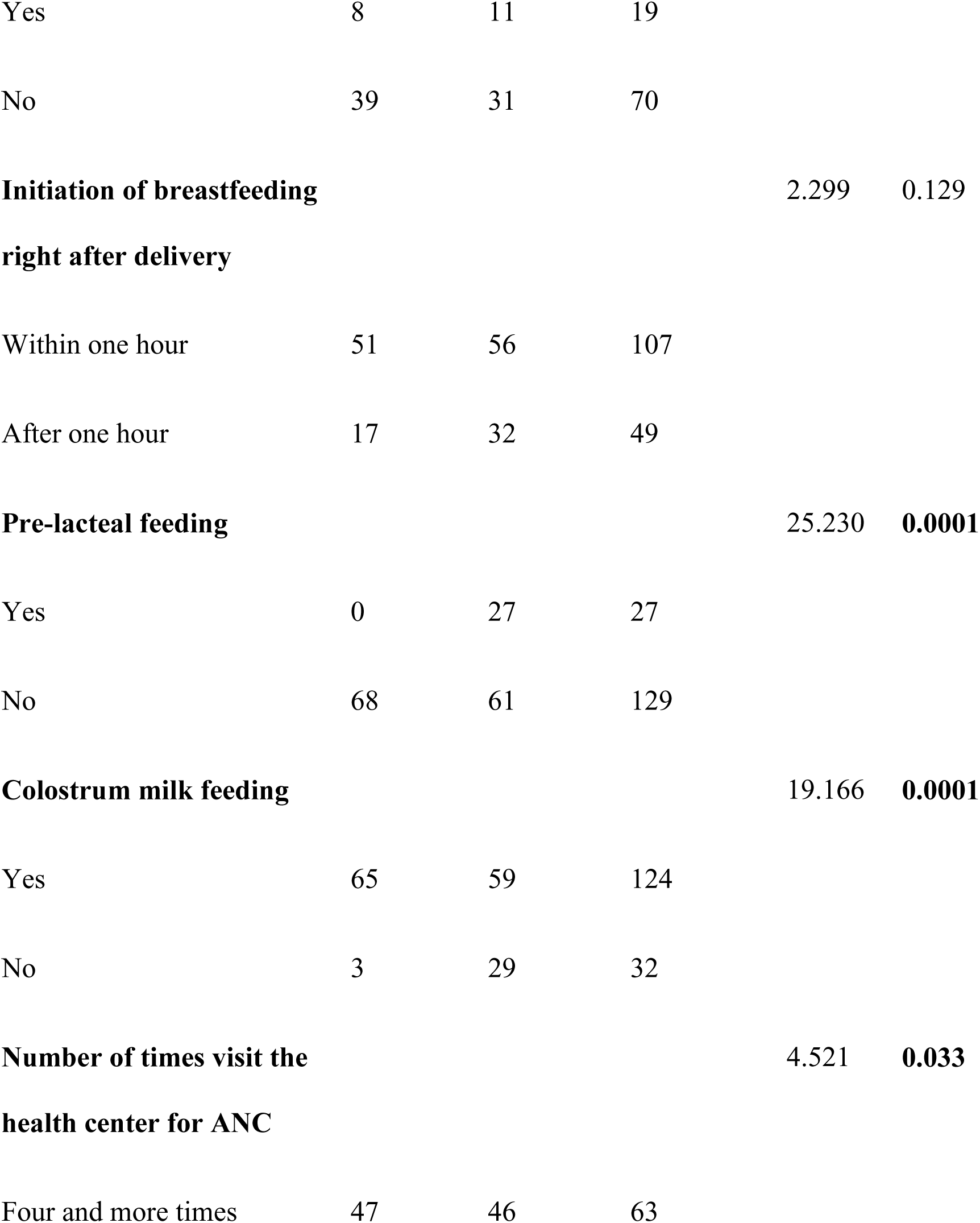
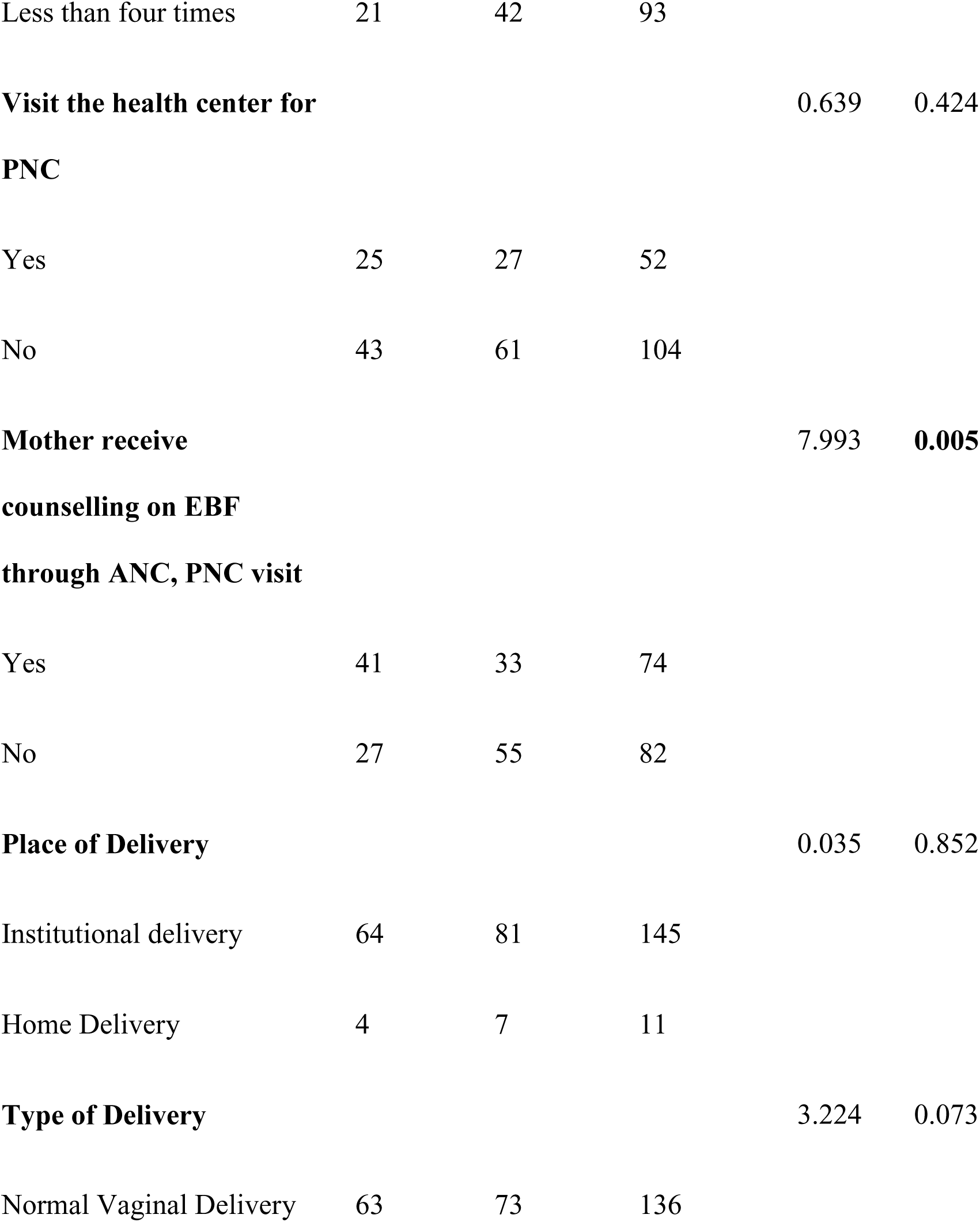
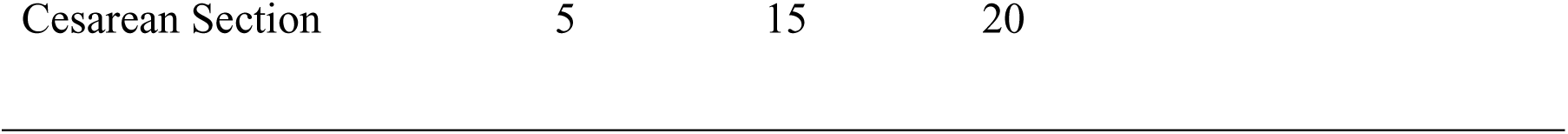
Association between other factors and exclusive breastfeeding.

| Variable | Exclusive breastfeeding for six months | | | $\chi^2$ | P-Value |
| --- | --- | --- | --- | --- | --- |
|  | Yes | No | Total |  |  |
| **Knowledge about BF** |  |  |  | 7.162 | **0.007** |
| Yes | 47 | 42 | 89 |  |  |
| No | 21 | 46 | 67 |  |  |
| **Knowledge related to the benefits of EBF** |  |  |  | 4.224 | **0.04** |
| Yes | 33 | 37 | 70 |  |  |
| No | 14 | 5 | 19 |  |  |
| **Knowledge related to EBF** |  |  |  | 7.371 | **0.007** |
| Yes | 40 | 25 | 65 |  |  |
| No | 7 | 17 | 24 |  |  |
| **Knowledge related to the management of Breast related problem** |  |  |  | 1.111 | 0.292 |
| Yes | 8 | 11 | 19 |  |  |
| No | 39 | 31 | 70 |  |  |
| **Initiation of breastfeeding**<br>**right after delivery** |  |  |  | 2.299 | 0.129 |
| Within one hour | 51 | 56 | 107 |  |  |
| After one hour | 17 | 32 | 49 |  |  |
| **Pre-lacteal feeding** |  |  |  | 25.230 | **0.0001** |
| Yes | 0 | 27 | 27 |  |  |
| No | 68 | 61 | 129 |  |  |
| **Colostrum milk feeding** |  |  |  | 19.166 | **0.0001** |
| Yes | 65 | 59 | 124 |  |  |
| No | 3 | 29 | 32 |  |  |
| **Number of times visit the**<br>**health center for ANC** |  |  |  | 4.521 | **0.033** |
| Four and more times | 47 | 46 | 63 |  |  |
| Less than four times | 21 | 42 | 93 |  |  |
| **Visit the health center for PNC** |  |  |  | 0.639 | 0.424 |
| Yes | 25 | 27 | 52 |  |  |
| No | 43 | 61 | 104 |  |  |
| **Mother receive counselling on EBF through ANC, PNC visit** |  |  |  | 7.993 | **0.005** |
| Yes | 41 | 33 | 74 |  |  |
| No | 27 | 55 | 82 |  |  |
| **Place of Delivery** |  |  |  | 0.035 | 0.852 |
| Institutional delivery | 64 | 81 | 145 |  |  |
| Home Delivery | 4 | 7 | 11 |  |  |
| **Type of Delivery** |  |  |  | 3.224 | 0.073 |
| Normal Vaginal Delivery | 63 | 73 | 136 |  |  |

|  |  |  |  |
| --- | --- | --- | --- |
| Cesarean Section | 5 | 15 | 20 |

### Multivariate Analysis

Table 11 presents logistic regression results exploring factors influencing exclusive breastfeeding among mothers of children under 2 years old. The logistic regression analysis identified several factors influencing exclusive breastfeeding practices among mothers of children under 2 years old. Initially, there appeared to be a gender disparity, with male children showing higher odds of exclusive breastfeeding, but this significance diminished after adjusting for other variables, indicating potential confounding. Homemaker mothers exhibited significantly higher odds of exclusive breastfeeding compared to employed mothers, a trend that strengthened after adjustment. Similarly, mothers living in nuclear families initially showed higher odds of exclusive breastfeeding, although this significance disappeared after adjusting for other variables. Non-smoking mothers were consistently more likely to exclusively breastfeed, even after adjustment. While factors like knowledge about breastfeeding and advice on exclusive breastfeeding showed significant associations in unadjusted analyses, they lost significance after adjustment, hinting at the influence of confounding variables. Notably, feeding colostrum milk and frequent ANC visits remained strongly associated with higher odds of exclusive breastfeeding even after adjustment. These findings underscore the multifaceted nature of exclusive breastfeeding determinants and the importance of tailored interventions to promote and support breastfeeding practices.

**Table 11:**
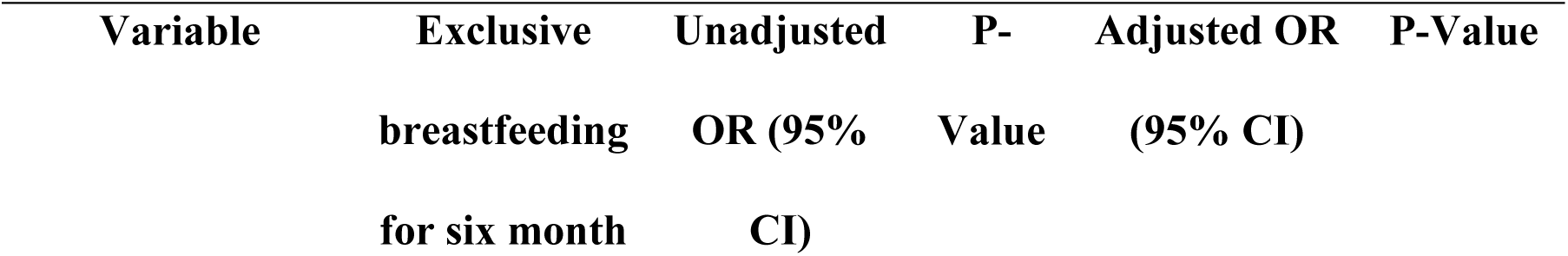
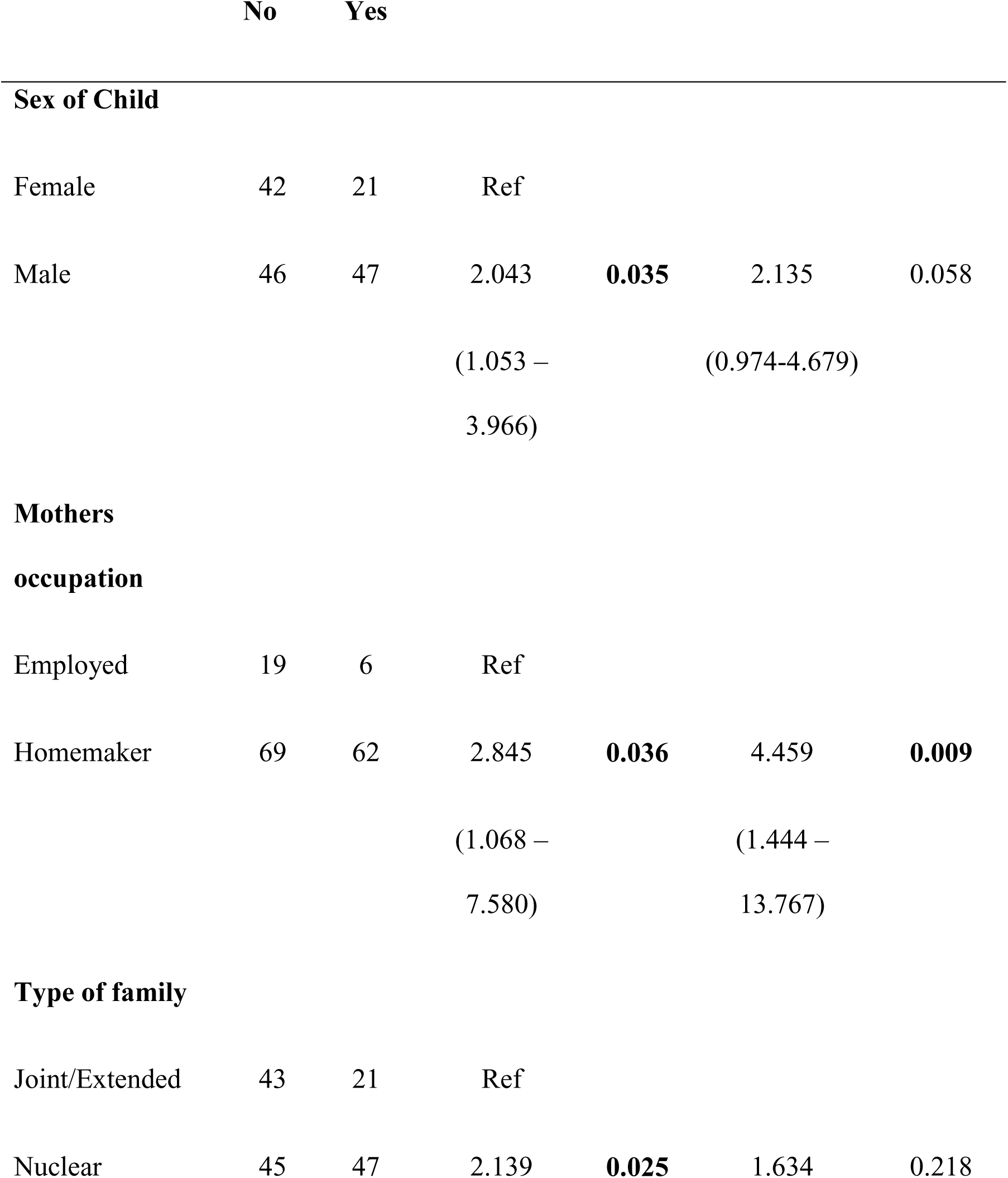
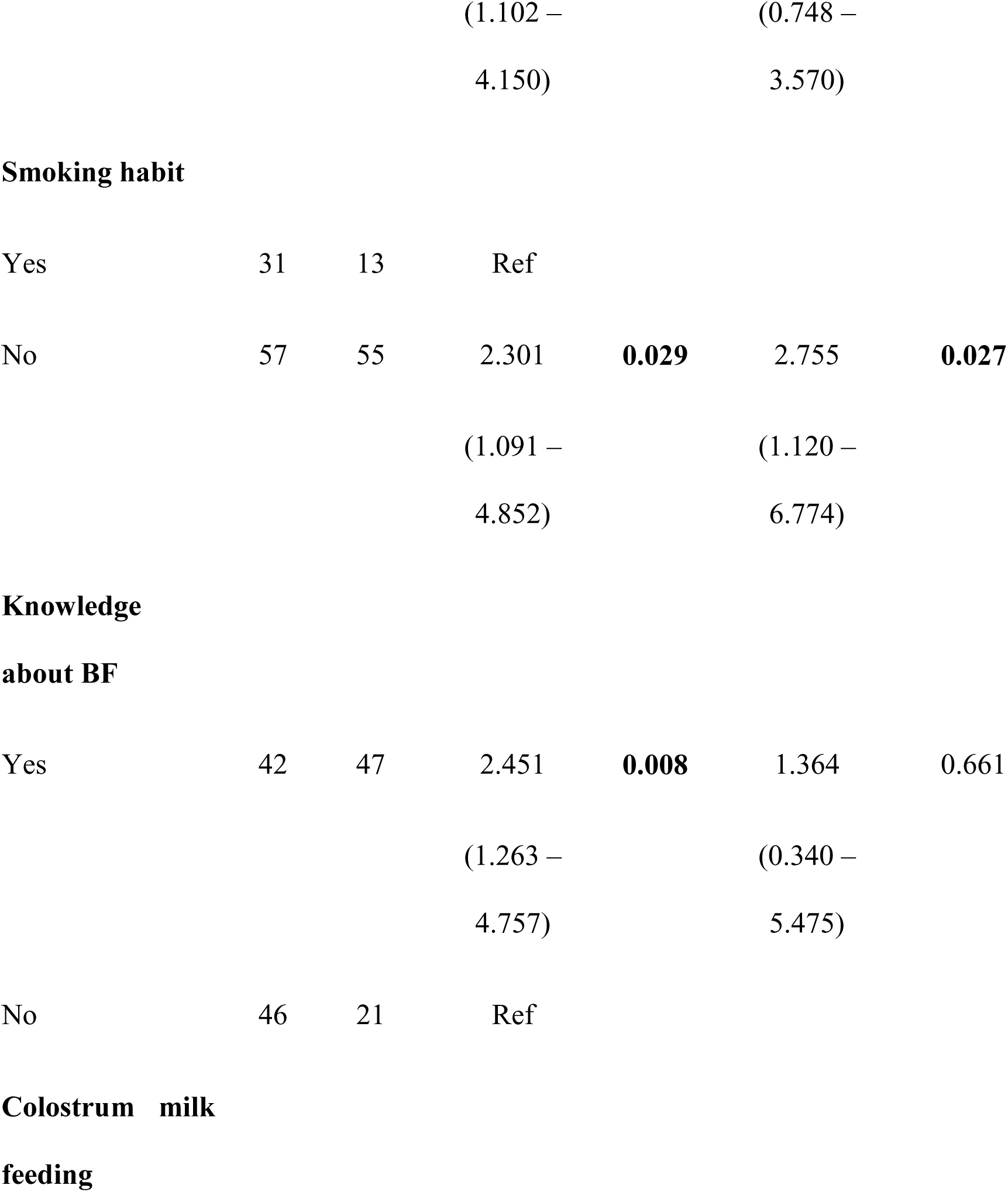
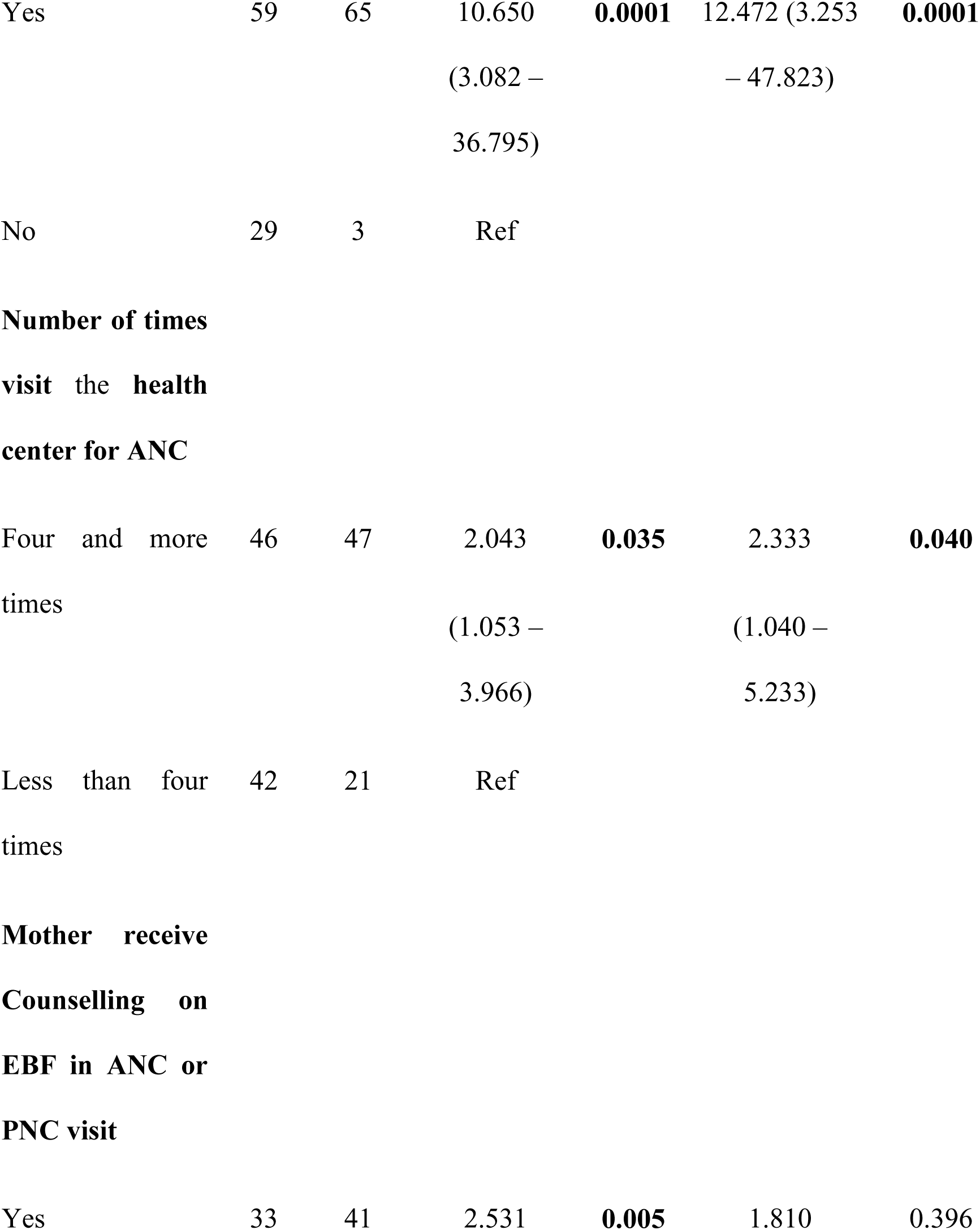
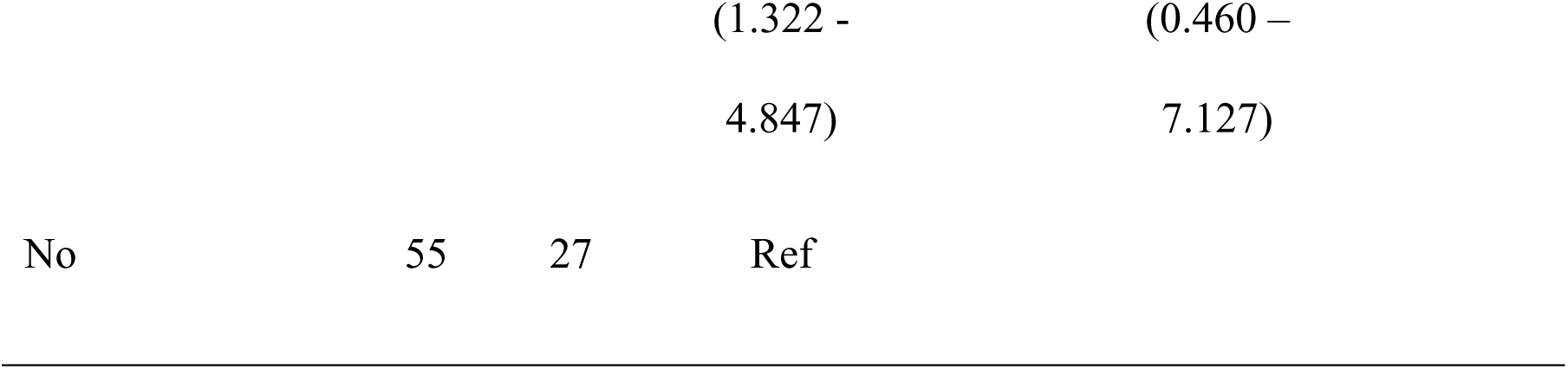
Multiple logistic regression.

## Discussion

An analytical cross-sectional study design was used to assess the proportion of exclusive breastfeeding and associated factors among 156 Mothers of children under 2 years of age in the Dalit community. This study aimed to assess the proportion of mothers of children under 2 years of age, practicing exclusive breastfeeding and associated factors in the Dalit community of Rajbiraj Municipality, Saptari, Nepal.

The study revealed an exclusive breastfeeding prevalence of 43.6%, showing a positive trend compared to the national average of 36.9%. Regional variations were observed, with Madhesh province at a lower 26%.(17) Discrepancies were noted in national reports, with NDHS 2022 at 56.4% and MICS 2019 at 62.1%.(18, 19) Comparable rates were found in western hilly region of Nepal reported a prevalence of 47.6%, (20) which was relatively similar to the our study. For instance, study conducted in the mid-western and eastern regions of Nepal found a rate of EBF 23.2%, (21) while Acharya et al. reported a prevalence of 53.0% in rural Terai of Nepal. (22) Dahal reported a prevalence of 49.1% in slum areas of Kathmandu Valley,(23) while Panthi found a rate of 71.5% among mothers in central Nepal. (24) (25) Sharma and Kafle reported that approximately nearly half (50.5%) in Pokhara Slum area, falling within our study’s range but precluding a direct comparison. (26) Globally, this findings align with the breastfeeding prevalence of (44%) but lower than the South-Asia region (57%). (27, 28) Comparing with neighboring countries show that present study has higher rate of EBF compared to India’s Bihar (27.6%), (29) and China (37%). The study presents encouraging trends in EBF prevalence within the Dalit community of Rajbiraj Municipality.

The current study observed that the age of the mothers ranged from 19 to 36, with a mean age of 24.59 years aligning with previous research in mid-eastern and western regions of Nepal and Western Nepal, where mean ages of 25.38 and 24.9 years were reported, respectively. (21, 30) Notably, studies conducted in the Slum of Pokhara and Nepalgunj emphasized a majority of participants being below the age of 30 year old. (29, 31) These studies complemented our findings, as their age ranges also matched those observed in our study. The association between the age of the mothers and exclusive breastfeeding practices was assessed using chi-squire statistics, which revealed no significant association (P = 0.079) in the current study. This result is consistent with the findings of study conducted in Rural Southern Nepal, another study conducted in Central Nepal and study in China reported no significant association between age and exclusive breastfeeding (P = 0.545, P = 0.12, and P = 0.73, respectively). (22, 24, 25) These consistent findings across different regions and countries suggest that age may not be a determining factor influencing exclusive breastfeeding practices among mothers in various contexts.

The findings of this study indicate that health workers have a substantial impact on promoting and influencing exclusive breastfeeding practices among mothers. The higher percentage of respondents being influenced by health workers (42.6%) compared to personal decision – making (35.3%) and influence from family/friends/community (22.1%) underscores the significance of health workers in EBF promotion. The comparison with the study conducted in Pokhara, further support this understanding, highlighting that (50.5%) of mothers receiving information on EBF from health workers. (32) the higher percentage of mothers influenced by health workers suggests their significant contribution to maternal decision-making, as they are trusted source of information capable of offering evidence based guidance. This emphasizes the importance of health worker engagement and education in promoting optimal infant feeding practices within communities.

The current study found that the child’s age in months ranged from 6 to 24 months, with a mean age of 11.39 months and a confidence interval of 10.64 to 12.14. The majority (64.1%) of the respondents had children in the 6 to 12-month age group, while 28.8% of the children were in the 13 to 18-month age group, and 7.1% were in the 19 to 24-month age group. Comparing these findings with the previous studies conducted in mid-western and eastern regions of Nepal, where the mean age of infants was 9.98±2.26 months, aligns with the child’s age range in the study. (21) A study in Central Nepal reported a median age of infants as 10 months (ranging from 4 to 14 months), falling within the observed age range (24) In the Slum of Pokhara, the distribution of children in the 6 to 12-month age group closely corresponds to the current study’s results. Additionally, also reported the distribution of children as 51.3% in the 6 to 12-month age group and 48.8% in the 13 to 24-month age group, which corresponds closely to the findings of the current study. (26) Similarly, a study in urban slums in Bihar, India, reported consistent distribution across age groups, albeit with slight variation in percentages, further supporting the reliability of the current study’s findings. (29)

The study, comprising 156 respondents, revealed that (59.6%) were male, and 40.4% were female. indicating a significant association with exclusive breastfeeding (P-Value = 0.003). specifically, male children had two-fold odds (OR=2.043) of being exclusively breastfed compared to female children, without adjusting for other factors (OR = 2.043; CI = 1.053 – 3.966). Comparing with previous studies, findings varied; rural southern Nepal reported 52.3% female children with no significant association with exclusive breastfeeding,(22) while study in Nepal and Dhulikhel Municipality found a slightly higher proportion of male infants, aligning with our results. (31, 33) In Central Nepal, the distribution was 58.8% male and 41.2% female, suffering from our study. (24) the slum of Pokhara reported 47% Male and 53% female infants, contrary to our findings. (26) In Pokhara more than half (57.4%) were male, aligning with our results, but they reported female infants more likely to be EBF (51.2%), contrary to our findings. (32) In urban slums, Bihar, India, the distribution was similar to our study, and they found that male infants had a higher chance of EBF for six months, supporting our findings. (29) these comparisons highlight the variability in gender distribution and its association with exclusive breastfeeding across different regions and studies. Our findings suggest a significant association between gender and EBF practices, emphasizing the need for further research to explore underlying factors influencing this relationship.

In the current study, out of the total 156 respondents, the majority (84%) were homemakers, while 16% were employed. The chi-squire analysis revealed a significant association between exclusive breastfeeding and mothers’ occupation (0.031). After adjusting for other variables, homemaker mothers had 5.724 times higher odds of practicing exclusive breastfeeding compared to employed mothers (OR = 2.845; CI = 1.068 – 7.580). These findings align with similar research conducted in Nepal. For instance, a study found that the majority (76.4%) of the mothers in their study were not formally employed, which aligns with the current study’s findings of a high proportion of homemakers. (33) Study conducted in central Nepal reported that a vast majority (74.3%) of the mothers in their study were homemakers, similar to the current study’s results. (24) Study conducted in Slum of Pokhara found that the occupation of mothers in theirs in their study was predominantly housewife (86.5%), which aligns with the current study’s results. (26) Study conducted in Urban Slum of India reported that almost 87.2% of the mothers in their study were housewives, which is consistent with the current study’s findings. However, they did not assess the association between mothers’ occupation and exclusive breastfeeding. (29) These consistent findings across different studies highlight the strong association between maternal occupation (homemaker vs. employed) and exclusive breastfeeding practices. The high prevalence of homemakers practicing exclusive breastfeeding underscores the potential impact of maternal employment status on infant feeding behaviors. Future research could further explore the underlying reasons for this association and identify strategies to support employed mothers in maintaining exclusive breastfeeding practices.

In the current study, approximately 68.6% of infants were fed breast milk within one hour right after delivery, while the remaining 31.4% were fed breast milk after one hour. The chi-squire analysis indicated no significant association between exclusive breastfeeding and the initiation of breastfeeding right after delivery (P = 0.129). Comparing these findings with National Data Annual Report (2077/78), Fifty-five percent (55%) of children age 0–23 months engaged in early initiation of breastfeeding. (17) The present study indicating a positive trend. Comparing these findings with the previous studies, there are mixed results regarding the association between the initiation of breastfeeding and exclusive breastfeeding. Some studies have reported a significant association, suggesting that early initiation of breastfeeding is associated with a higher likelihood of exclusive breastfeeding. (25, 26, 29) However, other studies, including the current study, did not find a significant association. (18, 24) It is important to note that the percentage of infants who were breastfed within one hour of delivery varied across the studies. While the current study reported 68.6%, other studies reported percentages ranging from 41.8% to 96.3%. (21, 31, 33) These variation could be due to differences in study populations, cultural practices, healthcare settings and other contextual factors. The timing of initiation of breastfeeding is an essential factor that can influence exclusive breastfeeding practices. While early initiation of breastfeeding is recommended and associated with positive outcomes, its direct impact on exclusive breastfeeding may vary depending on various factors unique to each study setting. Further research is needed to better understand the complex interplay between early initiation of breastfeeding and exclusive breastfeeding practices in diverse populations and settings.

The findings of the current study indicate that approximately four-fifths (79.5%) of the infants were fed colostrum milk, while 20.5% did not receive colostrum milk. The chi-square analysis revealed a significant association between exclusive breastfeeding and colostrum milk feeding in the study population (P = 0.0001). Furthermore, the study found that mothers who breastfed exclusively and fed colostrum milk had higher odds compared to mothers who did not feed colostrum milk, even after adjusting for other variables. Comparing these findings with the referenced data, it is evident that the prevalence of colostrum milk feeding varies across different studies. For instance, (21) reported a lower rate of infants not receiving colostrum (2.3%), while (33) found that 83.5% of infants were fed colostrum. (26)reported that 5.5% of mothers discarded colostrum milk before feeding breastmilk. (29) noted that the majority of mothers (78.5%) reported their babies receiving colostrum immediately after birth, while the rest were asked to discard it due to cultural beliefs. The relatively high prevalence of colostrum milk feeding in the study population (79.5%) is encouraging, as it indicates a positive trend toward optimal breastfeeding practice. However, the 20.5% of infants who did not receive colostrum milk highlights the need for targeted interventions and support to address this issue. Understanding the reasons behind this practice, such as cultural beliefs or lack of awareness, can help inform strategies to promote colostrum feeding and improve breastfeeding outcomes. Efforts to educate and empower mothers and families about the importance of colostrum milk and dispel myths or misconceptions surrounding its benefits are essential for promoting early and optimal breastfeeding practices.

The findings of the current study indicate that all respondents visited the health center for at least one-time antenatal care (ANC). Comparing with Annual Report 2021, Madhesh Province has the highest number 100% of adolescents who received first ANC services and First ANC visit, which aligns with this study. (17) NDHS (2022) Ninety-four percent (94%) of women reported receiving antenatal care from a skilled provider. (17, 18) These comparisons highlight the positive trend in ANC utilization observed in our study population, where all respondents sought antenatal care services at least once during their pregnancy. The high coverage of ANC visits is essential for promoting maternal and child health, ensuring timely interventions, and reducing maternal and neonatal mortality rates.

Among the total of 156 respondents, 59.6% visited the health center for ANC four or more times, while 40.4% visited the health center less than four times. The chi squire analysis revealed a significant association between exclusive breastfeeding and the number of times visiting the health center for ANC in the study population (P = 0.033). The odds of mothers who breastfeeding exclusively and visited a health facility for ANC four or more times were two-fold higher compared to those who visited less than four times (OR = 2.043; CI = 1.053 – 3.966). After adjusting for other variables, the odds increased to 2.333 (OR = 2.333; CI = 1.040 – 5.233). Comparing these findings with Annual report 2021, Four in five women (81%) had at least four ANC visits, this contradicts the findings. (17) Study conducted in Western Hilly region of Nepal reported that nearly half (49.7%) of the participants followed the recommended guidelines of attending ANC four or more times, which aligns with this study. (20) Study conducted in Pokhara observed that mothers who had ANC visits for four or more times had lower exclusive breastfeeding rates (47.6%) compared to those who visited for less than four times (57.9%). This contradicts the findings. (32) Urban slums, Bihar highlighted that completing three or more ANC visits was associated with prolonged exclusive breastfeeding (OR 151, 95% CI 21 – 537). (29) These comparisons highlight the variability in the association between ANC visits and exclusive breastfeeding across different studies and settings. While our study found a positive association between more ANC visits and higher rates of exclusive breastfeeding, other studies have reported contrasting results. Factors such as regional differences, cultural practices, and healthcare settings may contribute to these variations. Further research is needed to better understand the underlying mechanisms linking ANC utilization to exclusive breastfeeding outcomes and to tailor interventions accordingly to promote optimal infant feeding practices.

The findings of this study states that significant proportion of respondents in the study population did not receive post-natal care (PNC) services, with two thirds (66.7%) not accessing (PNC) services, and only one-third (33.3%) visited the health center for PNC services. The Annual Report FY 2077/78 (2021 years) reported that, proportion of mothers attending three PNC visits 25.1 percent and in Madhesh province 14.5%. The present study indicating a positive trend. (17) NDHS (2022) reported that, 70% of women received a postnatal check within the 2 days after delivery. this contradict the findings. (18) Additionally, study conducted in Western hilly region of Nepal reported that only 37.5% of participants went for postnatal services two or more times. This finding aligns with this study, where a significant proportion of respondents did not receive PNC services, indicating a potential gap in postnatal care provision. (20)

The current study indicates that almost half (47.4%) of the respondents received counselling related to EBF through ANC or PNC visits, while the remaining 52.6 % did not receive such counselling. The chi-squire statistics demonstrated a significant association between exclusive breastfeeding and mothers receiving counselling on exclusive breastfeeding in the study population (P = 0.005). Moreover, the unadjusted analysis revealed that mothers who breastfed exclusively and received counselling on exclusive breastfeeding through ANC or PNC visits had odds 2.531 times higher than those who did not receive such counselling (OR = 2.531; CI = 1.322 – 4.847). These findings align with previous research conducted in Nepalgunj Medical college, which indicated that only 59% of women received advice on breastfeeding during the antenatal period. (30) the study emphasized the importance of providing support and counselling routinely during ANC to prepare mothers for successful exclusive breastfeeding. Additionally, a study by Urban slum, Bihar reported that receiving counselling at the hospital was associated with higher odds of exclusive breastfeeding. this aligns with the present study’s results, highlighting the significance of receiving counselling on exclusive breastfeeding in promoting and establishing exclusive breastfeeding practices. (29) To enhance EBF rates, healthcare providers should prioritize the provision of evidence – based counselling and support throughout the antenatal, perinatal, and postnatal periods. This includes integrating counselling and education on exclusive breastfeeding into routine ANC, ensuring support is available during childbirth, and offering continuous guidance in the postnatal period to facilitate sustained exclusive breastfeeding. Strengthening health systems to prioritize and deliver quality counseling and support services is essential for improving exclusive breastfeeding practices and overall maternal and child health outcomes.

The findings of the study provided that among the total 156 respondents, 87.2% had a spontaneous vaginal delivery, while 12.8% underwent a cesarean section. The chi-square statistics showed no significant association between exclusive breastfeeding and the type of delivery in the study population, although it approached significance at the 10% level (P = 0.073). In the study conducted in Western hilly region of Nepal, the regression analysis revealed that exclusive breastfeeding was more likely among mothers who delivered through normal vaginal delivery. (20) Similarly, study conducted in Nepalgunj medical college found that mothers who had a normal delivery were two times more likely to practice exclusive breastfeeding compared to those who had an assisted delivery. (26) On the other hand, study conducted in Central Nepal reported that cesarean section was the most frequent cause (88.6%) for the late imitation of breastfeeding. (24) This suggests that cesarean section may have an impact on the timing of breastfeeding initiation, but its influence on exclusive breastfeeding practices may not be significant within the study population. The studies in Pokhara, Urban slum of Bihar, and China also assessed the association between mode or type of delivery and exclusive breastfeeding. (25, 29, 32) Study conducted in Pokhara found a significant association between mode of delivery and exclusive breastfeeding (p<0.001). (32) Urban slum of Bihar reported that mothers who delivered normally had two times higher odds of practicing exclusive breastfeeding compared to those who had a cesarean section. (29) In the present study, although the association between exclusive breastfeeding and type of delivery did not reach statistical significance, there is a trend suggesting a potential relationship. Further research with larger sample sizes and more detailed analysis may help clarify the impact of mode of delivery on exclusive breastfeeding practices in different populations and settings. Understanding these associations is crucial for developing targeted interventions to promote and support exclusive breastfeeding among mothers, regardless of their mode of delivery.

A significant proportion of respondents cited issues related to milk production, including trouble getting the milk flow to start, not having enough milk, or perceiving inadequate secretion of breast milk. These reasons were reported by multiple studies, including study conducted in Central Nepal,Nepalgunj medical college, and mid-western and eastern regions of Nepal. (21, 24, 30) Another common reason mentioned by respondents was the perception that breast milk did not satisfy the baby. This finding was reported in central Nepal (2022) and Nepalgunj medical college (2017). (24, 30) Some respondents reported specific difficulties with breastfeeding, such as painful breasts, difficulty in sucking and problems with latching. These challenged were highlighted in the studies in Central Nepal, and mid-western and eastern regions of Nepal. (21, 24) Several studies, including Nepalgunj medical college and Urban slum Bihar Mentioned household duties and work related problems as barriers to exclusive breastfeeding. (29, 30) Mothers may face challenges in balancing their household responsibilities and the time commitment required for breastfeeding. This can lead to decreased breastfeeding duration or early introduction of other foods or milk substituted. These findings highlight the multifaceted challenges faced by mothers in maintaining exclusive breastfeeding practices. Addressing these barriers requires comprehensive support systems, including targeted education and counseling, improved access to lactation support services, and workplace policies that accommodate breastfeeding mothers. Efforts to address these challenges are crucial for promoting and sustaining exclusive breastfeeding practices, which have significant benefits for maternal and child health outcomes.

### Conclusion

This study sheds light on the demographic characteristics, knowledge and breastfeeding practices among the Mothers of children under 2 years of age in Dalit community of Rajbiraj Municipality. The findings emphasize the need for targeted interventions and support to promote exclusive breastfeeding practices among mothers in the study population. Factors such as the sex of the child, the mothers’ occupation and education level and maternal knowledge were found to significantly influence exclusive breastfeeding. Adjusted analysis revealed that being a homemaker mother, completing formal education, feeding colostrum milk, and attending ANC visits frequently were significantly associated with higher odds of exclusive breastfeeding. However certain associations initially observed lost significance after adjusting for confounding variables, highlighting the importance of considering various factors and potential confounders in promoting exclusive breastfeeding.

### Recommendation

To promote exclusive breastfeeding among mothers and improve maternal and child health outcomes, the following recommendations are proposed:

1. Targeted interventions should focus on supporting breastfeeding practices especially among mothers who are employed or have lower levels of education.
2. Healthcare providers, particularly during antenatal care visits, should offer comprehensive and accurate information on exclusive breastfeeding, providing counselling and support to expectant and new mothers.
3. Identify and address common barriers and challenges encountered by mothers during exclusive breastfeeding, such as milk flow issues or concerns about infant satisfaction. Offer appropriate guidance, support, and interventions to overcome these challenges.

### Limitations

1. The study’s findings may not be generalizable to the entire population as it focused on a specific region or community.
2. Another limitation of this study was recall bias due to the retrospective nature of the data collection, possibly resulting in over or underestimation of actual feeding practices. Although recall biases cannot be avoided, the researcher conducted all interviews by asking probing questions to gather exact information.

## Supporting information

Informed Consent forms, Data collection tools, Approval letter from IRC

## Data Availability

All data produced in the present work are contained in the manuscript

