## Supplementary material for "Factors associated with exclusive breastfeeding among mothers of children under two years of age in Dalit community, Rajbiraj Municipality, Saptari, Nepal": Informed Consent forms, Data collection tools, Approval letter from IRC: Annexures.docx

Annex I- Informed Consent forms

Annex II- Data collection tools

Annex III- Approval letter from Institutional Review Committee of Nobel College

### Annex I: Informed Written Consent Form

Information sheet

Department of Public Health

Nobel College, Sinamangal

**Study Title:** Factor Associated with Exclusive Breastfeeding among mothers of children under 2-year age in Dalit Community, Rajbiraj Municipality, Saptari, Nepal

**Investigators:** Neha Kumari Das

Address: Nobel College, Sinamangal

Phone no: 9803031137

**Background and Purpose:** My name is Neha Kumari Das. I am studying master of Public Health at Nobel College, Sinamangal. I am going to conduct this research to access “factors associated with exclusive breastfeeding among mothers of children under 2 years of age in the Dalit community, Rajbiraj Municipality, Saptari, Nepal” as a partial fulfillment of our curriculum. The information gathered from this study was used for study purposes. I hope that findings from this study can be supported to address the current situation and associated factors of exclusive breastfeeding among mothers of infants.

**Risks and Benefits:** You are selected for voluntary participation in research. Before you decide to participate in this research, it is important to understand that you will not benefit directly from it right now, but the findings from this study may help you benefit in the long run if any government or nongovernmental organization launches an exclusive breastfeeding program based on the findings of this study. And this study may also help policymakers develop new policies related to exclusive breastfeeding which help to improve the health of mother and child. There is no risk of participating in this study.

**Procedure:** We will interview you in a secure location so that no one can overhear our discussion. You will be given answers regarding demographic, and exclusive breastfeeding-related questionnaires. It will take 15-20 minutes to answer all these questions.

**Confidentiality:** The information you tell us will be used only for this study, and information from this interview may be presented at professional meetings or in written articles, your name will not be used and not identified anywhere in the questionnaire, reports of other publications, and presentations. All information given will be kept private and confidential. We will keep the information secure and in a safe place.

**Withdrawal of participation:** In case you don't feel comfortable with the questions or you don't like to participate in the discussion, you will be free to withdraw at any time during the study, without giving any reason.

**Payment:** We will not pay you for your participation.

If you have any question about this study, you may contact the following people.

Name: Neha Kumari Das

Mobile Number: 9803031137

Nobel College, Sinamangal Kathmandu

**Are you willing to participate, kindly give me your consent?**

**Willing to take part in study	Yes	No**

**Factor Associated with Exclusive breastfeeding among Mothers of children under two year of age in Dalit Community, Rajbiraj Municipality, Saptari, Nepal**

Code No:

Namaste!

We are students of "The Master of Public Health (MPH)" at Nobel College, Pokhara University. We are conducting research on "FACTOR ASSOCIATED WITH EXCLUSIVE BREASTFEEDING AMONG MOTHERS OF CHILDREN UNDER TWO YEARS OF AGE IN THE DALIT COMMUNITY, RAJBIRAJ MUNICIPALITY, SAPTARI NEPAL" as a partial fulfillment of our curriculum. The information gathered from this study will be used for study purposes and will be kept confidential. We assure you that there are no risks for you in taking part in this study. In this context, we would like to ask you some questions about factors associated with exclusive breastfeeding among mothers of children under two years of age in the Dalit community, of Rajbiraj Municipality, Saptari, Nepal. It will take about 20–25 minutes. Your participation in this study is voluntary. You are free to withdraw from the interview at any time or refuse to answer any particular question that makes you feel uncomfortable. We would appreciate it if you participated in the study and answered all the questions, as the information you provided would be very important for this study. There is no harm in you being a part of the study.

…………………………	………………………

Signature of respondent	Date

### Annex II- Data collection tools

**QUESTIONNAIRE**

**Survey information**

| **Location and date** \| \| | | **Response** |
| --- | --- | --- |
| 101 | Ward Number |  |
| 102 | Date |  |
| **Consent and interview name** | | **Response** |
| 103 | Consent has been heard, read and obtained ? | Yes \| 1 \|; No \| 0 \| |
| 104 | Family surname |  |

**Demographic Information**

| **Demographic Information** | | |  |
| --- | --- | --- | --- |
| **Question** | | **Response** | **Code** |
| 201 | How old are you? | ……………… year |  |
| 202 | How old your child? |  |  |
| 203 | Sex of child | Male  Female | 1  2 |

| 204 | What is your occupation? | Employed  Homemakers | 1  2 |
| --- | --- | --- | --- |
| 205 | What is your husband occupation? | Private or government worker  Labor Worker  Un employed/self employed  Foreign Worker | 1  2  3  4 |
| 206 | What is your education status? | Cannot read and write  Read and Write  Primary education  Secondary education and above | 1  2  3  4 |
| 207 | What is your husband education status? | Cannot read and write  Read and Write  Primary education  Secondary education and above | 1  2  3  4 |
| 208 | What is your type of family? | Extended /Joint  Nuclear | 1  2 |
| 209 | How many children do you have? | One  Two  Three and above | 1  2  3 |
| 210 | Do you have smoking habit? | Yes  No | 1  0 |
| 211 | Do you drink alcohol? | Yes  No | 1  0 |

**Breastfeeding related Information:**

| **S.N** | **Question** | **Response** | **Code** |
| --- | --- | --- | --- |
| 301 | Did you hear about breastfeeding? | Yes  No (If no then skip Q. No. 302, 303) | 1  0 |
| 302 | If yes, what did you hear about breast feeding? | Benefits of breast feeding  Exclusive breastfeeding  Management of Breast related problem |  |
| 303 | Who gave you the information about exclusive breastfeeding? | Health Worker  Friends/Community  Social Media/FM/Redio/TV  Meeting of Mothers group |  |
| 304 | When did you start breast feeding right after delivery? | Within one hour  After one hour | 1  2 |
| 305 | After delivery, did you feed your baby anything before breastfeeding? (Pre-lacteal feeding) | Yes  No (If no then skip Q. No. 309, and 310) | 1  0 |
| 306 | Did you feed your baby colostrum milk? | Yes  No | 1  0 |
| 307  (Dep.V) | Did you breastfeed your baby exclusively for six months? | Yes  No (Skip Q. No. 309) | 1  0 |
| 308 | When did you first introduce anything solid or liquid food except breastmilk to your child? | 0 to 2 Months  3 to 5 Months  6 Month and above | 1  2  3  4 |
| 309 | Who influenced or advice you to feed breast milk exclusively? | Friends/family/Community  Health worker  My own decision | 1  2  3 |
| **310** | **What are the following reasons for your decision to stop breastfeeding your baby?** | My baby had trouble sucking or latching  My baby become sick and could not breastfeeding  Breast milk not satisfy my baby  I thought my baby was not gaining enough weight  I had trouble getting the milk flow to start  I didn’t have enough milk  I had too many household duties |  |
| 311 | Did you visit health center for antenatal (ANC) checkup? | Yes  No (If no then skip Q. No. 319, 320) | 1  0 |
| 312 | How many times did you visit health center for the ANC. | Less than four time  Four and more time | 1  2 |
| 313 | Did you visit health center for post-natal care? | Yes  No (if no then skip Q. No. 324, 325) | 1  0 |
| 314 | Was the mother receive counselling on exclusive breastfeeding through ANC,PNC visit | Yes  No | 1  0 |
| 315 | Where did you gave birth to your child? | Institutional Delivery  At home | 1  0 |
| 316 | What type of delivery did you have? | Normal vaginal delivery  Caesarean section | 1  0 |

**Annex III- Approval letter from Institutional Review Committee of Nobel College**

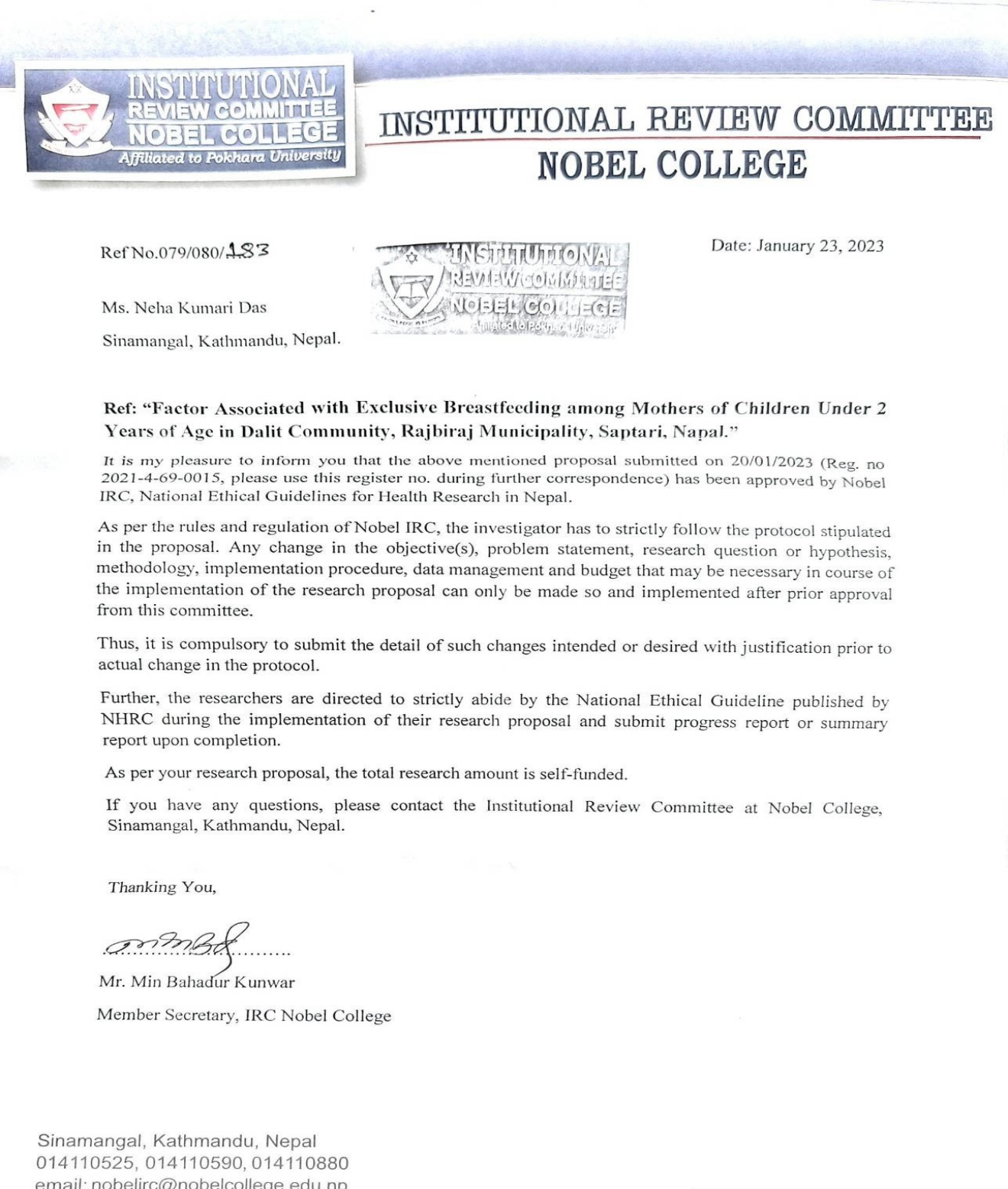
